# Modelling learning trajectories of facial emotion recognition training in autistic children

**DOI:** 10.1101/2025.09.29.25336883

**Authors:** Osama Mahmoud, Dejla Hoxha, Ian S Penton-Voak, Christopher Jarrold, Marcus R Munafò, Angela S Attwood, Zoe E Reed

## Abstract

**Background:** Emotion recognition (ER) skills are important in social interaction. ER difficulties are associated with social difficulties and may lead to poorer educational outcomes and wellbeing. There is therefore considerable interest in tools that support development of ER skills. In addition, given that autism is associated with ER difficulties, such tools may be particularly beneficial for autistic children. In this study we examined how autistic children’s ER skills changed over 8 sessions of using an ER training task.

**Methods:** Autistic schoolchildren or those considered to be autistic by parents (N=88; 79 with complete data) were recruited from school and community settings to complete 8 sessions of the digital ER task. We used mixed-effects regression models to estimate learning growth across all 8 sessions. We also examined how participant characteristics influenced results. Finally, we identified the optimal number of sessions needed for this task in terms of ER improvement.

**Results:** We found overall improvements in ER with mean accuracy scores increasing from 58 to 73 (out of 100), representing a 25% improvement. Age, verbal age, gender, parental education and income influenced improvements in ER. The optimal number of sessions was 6.

**Conclusions:** Our study demonstrated that completing multiple sessions of the ER training task was associated with improvements in ER. Improvements were seen across all children, but some characteristics influenced the pattern and degree of this change. Our study utilised mixed-effects smoothing spline models, bringing novel value to this context. These findings support the view that ER is a tractable target and inform how future interventions/tools may be implemented.

## Introduction

Emotion recognition (ER) skills are an important part of social interaction, and difficulties in recognising other people’s emotions can have a substantial negative impact on social interaction and relationships ^1–3^. For example, experiencing more ER difficulties is associated with having more peer problems, social problems, and social withdrawal. Additionally, ER difficulties may negatively influence academic achievement^3,4^ and mental health^5,6^. Consistent with this, some ER training tools/interventions appear to improve downstream outcomes such as mental health^7^. ER skills tend to develop during childhood and adolescence, with ER ability increasing with age^8–10^ and therefore difficulties with ER likely emerge during this period. These difficulties are more likely to be experienced by neurodiverse children, in particular autistic children or those with autistic traits^11–14^. Given this, any intervention supporting the development of ER skills, particularly during childhood and adolescence, is likely to have the most benefit on downstream outcomes.

There are several evidence-based tools currently available that aim to support development of ER skills, often targeting ER difficulties in autistic individuals^15–21^. Many (although not all) focus on facial ER. These tools tend to be digital and tend to focus on the emotions of happiness, anger, sadness, fear, surprise and disgust. Meta-analytic results have demonstrated that ER is a tractable target, in particular for autistic individuals^15,16^. We recently developed a facial ER training task which includes different intensities of emotional expressions for all six of the basic emotions has demonstrated improved ER in the general population and autistic adults, as well as generalisability beyond training stimuli^22,23^. This new training task goes beyond existing tools by presenting stimuli with emotional expressions at a range of intensities, from more subtle to more pronounced, providing varying levels of difficulty. It can also be accessed both in-person and online with minimal supervision. Importantly, this task is part of a wider toolkit that is being co-designed with autistic children and adults, parents and school staff, ensuring that it is relevant and acceptable to the autistic community. Many existing tools are limited by their evidence-base, lack of accessibility (i.e., only available offline or lack of ability to tailor (e.g., adjusting difficulty levels).

There is also limited information on how well these ER tools support children over time (i.e., across repeated sessions). In the present study we examined whether there were improvements in ER in autistic school aged children, (where some children had a formal diagnosis and others did not, but were considered to be autistic by parents or were on the waiting list for a referral) after using this task over 8 sessions.

The specific aims of this study were to:

1. Examine whether a recently developed ER training task steadily improves ER in autistic children across repeated sessions
2. Understand how learning growth varies based on different characteristics of autistic children
3. Identify the optimal number of sessions (identified as a tapering off or plateauing of the learning growth curve)

In addition, we compared different statistical models to understand patterns of learning over time, as the degree of growth (i.e., increases in accuracy from one session to the next) may not be linear across all of the sessions. Understanding this pattern of learning, and how it may differ by participant characteristics, can inform future implementation, whereby the number of sessions required could be tailored to the individual.

## Methods

The protocol for this study was pre-registered on the Open Science Framework (https://doi.org/10.17605/OSF.IO/429EM). Full details on deviations from the protocol are outlined in Supplementary Materials, Section 1.

### Data and code availability

The data and analysis code that form the basis of the results presented here are available from the University of Bristol’s Research Data Repository (http://data.bris.ac.uk/data/), DOI: available on publication).

Ethics approval was obtained from the School of Psychological Science Research Ethics Committee at the University of Bristol (Reference: 10493). Informed consent was obtained from parents of participants through a parental survey in Qualtrics after being presented with the information sheet. Parents were provided with contact details of the researcher should they wish to contact them with any questions or concerns before continuing. Parents were asked to indicate in the online parental survey whether they consented for them and their child to take part or not. They were presented with the statement “I hereby fully and freely consent to my and my child’s participation in this study as detailed above” after viewing the information sheet and consent form details. Participants could select “I consent” or “I do not consent”. Following parental consent, researchers met with children at their school in one-to-one sessions. The researcher explained the study to the child, then the child was asked to provide their verbal assent to take part in the study.

#### Study design

Participants completed up to 8 sessions of facial emotion recognition training (ERT), and longitudinal modelling was used to examine changes in ER accuracy over these sessions. The primary outcome measure was total hits (i.e., accuracy as identified by the number of correct identifications of emotional faces) in each training session.

#### Recruitment and participants

Parents of autistic children were initially recruited via schools. In response to slow recruitment rates, two additional approaches were later introduced: via community groups/centres and local advertisements, and via social media.

##### School approach

Special educational needs (SEN) and mainstream schools within Bristol, UK, were contacted by email, telephone, or in-person visits by the researchers. Interested schools were emailed an information sheet explaining their role in the study and an invitation to send to parents asking them if they would be happy for their child to take part in the study. Schools invited researchers to introduce the study to students during assembly time and forwarded the study invitation to parents via school newsletters, individual and group emails, and/or printed letters sent home with the students. This invitation provided basic study information and a link to a Qualtrics survey (www.qualtrics.com), which presented the full information sheet and contact information of the researchers. Parents used this link to consent and sign their child up to the study as well as to complete a parental survey (see “Parental Survey” below for details).

Following parental consent, researchers met with children at their school in one-to-one sessions. The researcher explained the study to the child, then the child was asked to provide their verbal assent and agreed to meet for a total of 8 sessions to complete the ER task. Children were free to withdraw from the study at any time.

To thank children for their participation, they received a certificate of recognition as well as a gift-bag or a shopping voucher (children selected which they preferred). The voucher/gift-bag was valued at £5 for children who completed 3-5 training sessions and £10 for children who completed 6-8 sessions. Parents received £10 for completing the parental survey. Schools were offered a token of thanks for their support and time, which comprised a £50 enrolment fee regardless of the number of children that participated from that school and an additional £10 per child that completed at least six sessions.

##### Community group approach

The study was advertised to parents through local centres/groups and other advertisement means (i.e., study posters displayed on noticeboards around the University of Bristol campus and local public spaces, using similar invitations and information sheets to those recruited via schools). Most community recruitment was done through adventure playgrounds around Bristol, UK. These are recreational spaces where children and young people engage in unrestricted play during organised or free sessions, with the support of playworkers. Many adventure playgrounds offer dedicated hours and support for neurodiverse children. Consent and reimbursement for community sites and community-recruited children was the same as the school approach (see above).

##### Social media approach

An advertisement for the study was shared on Facebook social media groups (i.e., local Bristol Facebook groups). Interested parents contacted the researchers directly via the contact details in the advertisement. Additional information and the link to the Qualtrics survey was forwarded to these parents. Participation via the social media route required additional commitment from the parents compared to participation via schools (i.e., additional parent time for arranging and facilitating sessions compared to researcher only facilitation; sessions taking from free time after school and work compared to sessions taking place during school time; travel time and cost if sessions were held in-person). Therefore, parent reimbursement was increased to £20 (shopping voucher) and travel was reimbursed as needed.

To be eligible children needed to be aged 7 to 16 years, fluent in English, have received a diagnosis of autism or considered to be on the autism spectrum by parents/carers (e.g., due to long waiting times for assessment some children were in this process and did not yet have a diagnosis, or the schools/parents considered them to be potentially autistic), and not have an uncorrected visual impairment (all parent reported). A further eligibility criterion was that the child must not have a verbal age lower than 5 according to the British Picture Vocabulary Scale (BPVS) (administered by researcher). The latter was assessed in the first session with the child after parents competed the survey and consented for their child to take part but before any ER tasks. Children were given an information sheet (there were two shorter child friendly versions, one for children aged 7 to 12 and one for children aged 13 to 16) about the study which they could read themselves or have the researcher read through with them. They were asked whether they wanted to take part (i.e., assent obtained) before the first session commenced and asked if they were happy to continue at the start of each subsequent session.

The target sample size was determined using the ‘Power IN Two-level designs’ (PINT) software^24^. We estimated that a sample size of n=110 children and at least six sessions were sufficient to be 95% confident in detecting an effect size, i.e., rate of change (learning growth) of at least 5%, with a power of 90% and a standard error (precision metric) of 1%. Our sample size calculations were based on preliminary data collected by our team in previous unpublished work, from a sample of 12 children with four repeated measurements. The statistical inputs for the ‘PINT’ software were informed by our analysis of this data, where the estimated variances of child’s averages and learning growth rates were 3.1% and 0.3% respectively, with an association (covariance) between the two measures of -0.009, and an estimated residual variance of 0.4%. We conducted up to eight sessions with children, but the main analysis included data from any child who completed at least six sessions. Any child that completed five or fewer sessions was replaced for the purpose of the main analyses. However, their data were retained for exploratory analyses. Our final sample size of children completing at least six sessions was 88 (79 completing all eight sessions). This was less than the target sample size due to difficulties with recruitment. However, this sample size was sufficient for our analyses, albeit with a reduced power (83%) and precision (standard error = 1.8%) to detect the same effect size mentioned above. 79 children (90%) completed all 8 sessions, providing a complete-case dataset with sufficient size (power = 79%; standard error = 1.9%). The other 9 children completed fewer than 6 sessions. Attendance at sessions was high overall (92.5%) with a total number of 651 sessions completed out of a possible 704. Recruitment and study completion are outlined in Figure 1.

**Figure 1.**
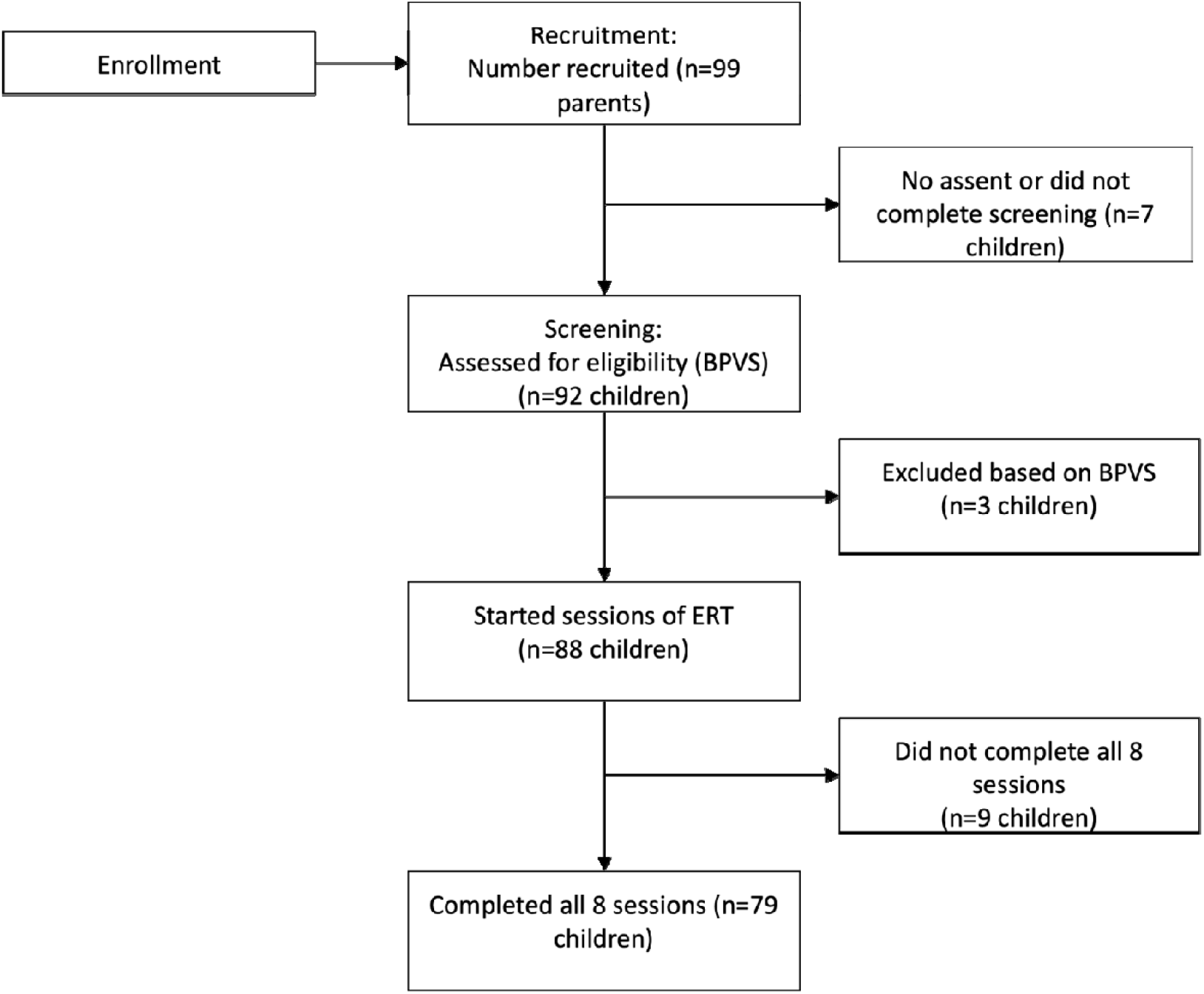
CONSORT flow diagram. ERT=emotion recognition training, BPVS= British Picture Vocabulary Scale

#### Study procedure

The training sessions were completed by participants in the presence of a researcher either in person or online. Per protocol, sessions were at least 24 hours apart. We stipulated that all sessions would be completed across 4-weeks (2 per week) where possible, but that exact timings would be at the discretion of the school and dependent on the availability of children with an aim of an upper limit of 8 weeks. In fact, all 8 sessions were completed within 13 to 111 days, mean 41 days (SD=22 days). The large range in days was due to a combination of school absences of children, school holidays, and logistics of session arrangement. Where the time taken to complete sessions was shorter than expected (i.e., more than 2 sessions per week) this was similarly due to logistics of arranging sessions within the school term time.

#### Parental survey

We collected information about the participating parent and their child via an online survey using Qualtrics software. This survey collected demographic data (i.e., child gender, child age, parental education, household income) and information on any child diagnoses of learning disability, attention deficit hyperactivity disorder (ADHD), developmental language disorder (DLD), dyslexia and dyspraxia (see Table 1 for full details of the information collected and how these were used in analyses). In addition, child autistic traits were measured through the parent-completed Child Autism spectrum Quotient 50 (AQ50)^25^ and child alexithymia was measured through the parent-completed Children’s Alexithymia Questionnaire (CAM)^26^.

**Table 1.**
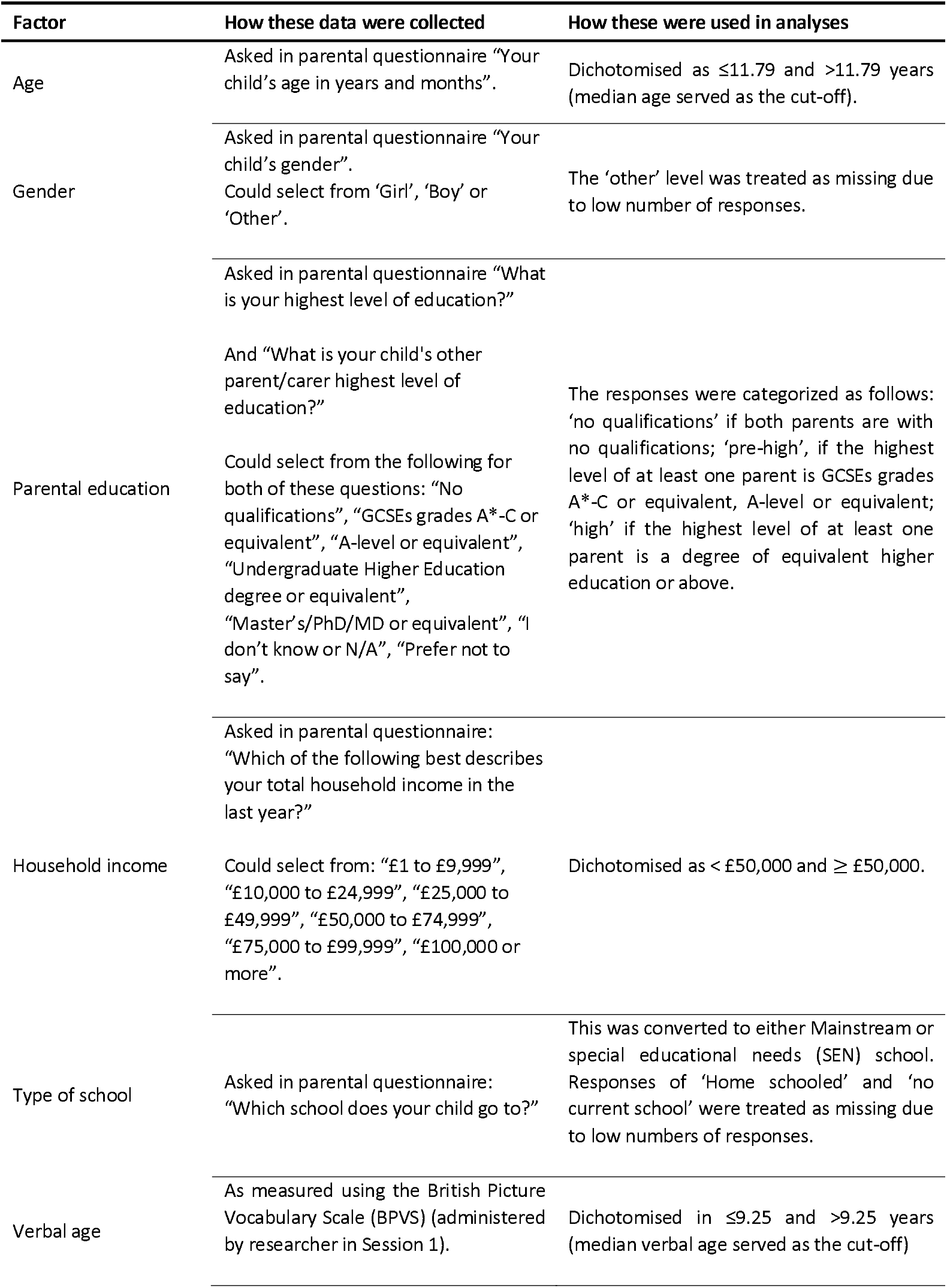
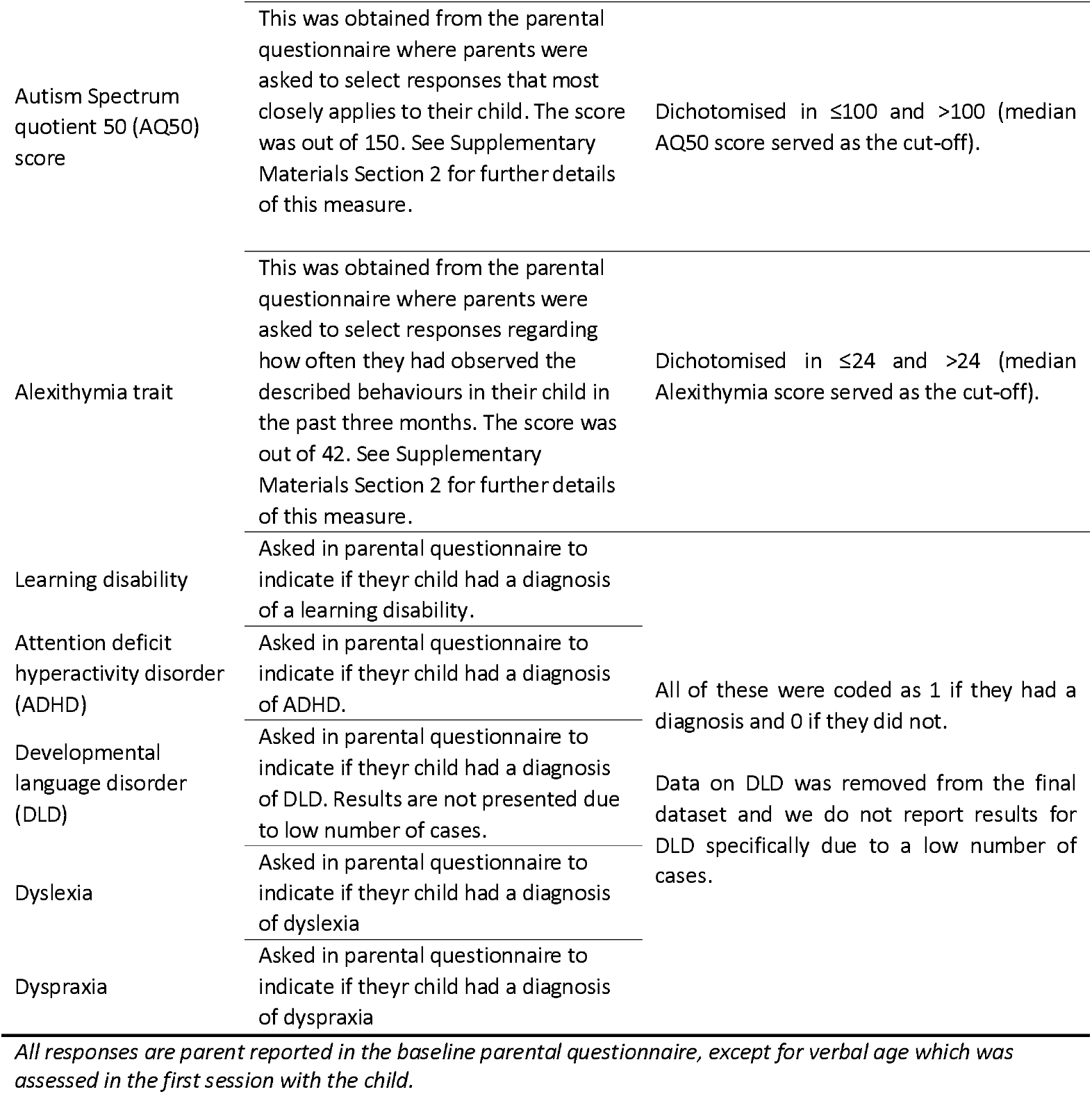
Description of variables collected through the parental questionnaire and from the child in Session 1. Variables relate to the child specifically unless otherwise specified.

##### Child sessions

Sessions were either completed in person in school, in person outside of school (i.e., at the University of Bristol), or online on a video call with the researcher, depending on parent and child preferences, if outside of school. For in-person sessions outside of school, parents were able to attend our research facility (at University of Bristol).

Session 1 involved sharing an information sheet with the child. Once assent was provided and recorded, the researcher completed the BPVS with the child. If they met the BPVS eligibility criterion of a verbal age higher than 5, they moved onto the ERT (see below). If the child did not meet this eligibility criteria, they were informed that they were not able to take part in the full study and thanked for their time. The first session took between 20 to 60 minutes, varying across participants. In each of the following sessions (session 2 to 8) the child was asked if they were happy to continue to take part at the beginning of each session before proceeding with the ERT. Each session lasted between 5 and 20 minutes, varying across participants. At the end of session 8, the child was asked about their experience of the ERT (see Supplementary Materials Section 4). Finally, they were presented with the study debrief, which was also sent to parents, and a giftbag or voucher.

##### Emotion recognition training

The ERT was hosted on the Gorilla platform (https://gorilla.sc/).

In the ERT, participants were presented with images of faces expressing one of six emotions (happy, angry, sad, scared, surprised, and disgusted) at one of 8 levels of intensity, ranging from a mild expression of the emotion to a more pronounced expression of the emotion. Facial stimuli were realistic-looking computer-generated images amalgamated from photos of 12 real-life individuals expressing these emotions and are therefore not identifiable^27^. Participants were presented with all 48 images during a session. A fixation cross was presented for 800 milliseconds (ms) before stimuli were presented on screen (either on an iPad if in person/at school or on a suitable device at home, if online, although mobile phones could not be used due to the task design) for 1000 ms, then masked for 250 ms. Then six emotion words corresponding to the six facial emotions were displayed. Participants were asked to select the emotion they thought they had seen. If they guessed incorrectly, they were presented with the face again and asked to try again until a correct response was given (maximum of 6 tries). Further details on the task can be found in our previous publication^22^. Accuracy scores (described below) were derived for each participant at each session from the total hits across all emotions (48 facial images). Before completing the ERT for the first time, children completed a practice task showing images of fruit and vegetables with corresponding words, to ensure understanding.

##### Statistical analyses

Descriptive statistics of the characteristics for the whole study population (n=88), participants who completed all eight sessions (n=79) and those who did not complete all 8 sessions (n=9) are presented in Table 2.

**Table 2.** Characteristics of study population (n = 88), participants who have attended all the eight sessions (n = 79), and those who have attended less than eight sessions (n = 9)

| Factor | % or median |  |  |
| --- | --- | --- | --- |
|  | All participants | Attended all 8 sessions | Attended < 8 sessions |
| Age (years)* | 11.79 | 11.75 | 13.67 |
| Female | 42.53 | 42.31 | 44.44 |
| High parental education | 46.59 | 46.84 | 44.44 |
| Pre-high parental education | 34.09 | 34.18 | 33.33 |
| Household income ≥ £50k | 39.47 | 41.18 | 25.00 |
| SEN school | 44.71 | 44.74 | 44.44 |
| Verbal age (years)* | 9.25 | 9.25 | 10.04 |
| Verbal age – age (years)* | -0.75 | -0.75 | -1.13 |
| AQ50 score* | 100.00 | 100.00 | 100.00 |
| Alexithymia score* | 24.00 | 24.00 | 28.00 |
| Learning disability | 19.32 | 17.72 | 33.33 |
| ADHD | 26.14 | 24.05 | 44.44 |
| Dyslexia | 6.82 | 7.59 | 0.00 |
| Dyspraxia | 9.09 | 8.86 | 11.11 |

\*Continuous scale where medians were calculated

To increase power and minimise selection bias, we performed our primary statistical analyses on data from the whole study population. In our secondary analyses, we repeated the analyses using data from participants who completed all eight sessions only (complete-case analyses).

For each participant, the accuracy score achieved during a single session (*Li*(*i* = 1, 2, …, 8) was defined as percentage correct hits in that session (see Supplementary Materials Section 5). Longitudinal measurements of accuracy (referred to as learning trajectories) were the primary outcome in our statistical analyses.

Three longitudinal models with different structures were fitted to estimate the learning trajectories across sessions (S1 to S8) each designed to capture different levels of complexity and underlying patterns in the data. Supplementary Table S1 details the different models used. These models, referred to as ‘M1: Main’, ‘M2: Monotonic’, and ‘M3: Reduced’, are structured as follows. M1 provides a flexible and detailed approach, where smooth cubic functions are employed to model both the overall trend in learning (i.e., shared across all children) and individual differences (i.e., how each child’s learning might vary). M1 can then capture not only how learning changes over time on average, but also how each child’s onset and learning trajectory may differ from that of other children in more complex ways. M2 has similar specifications to M1, except that it processes monotonic non-decreasing patterns of learning, i.e., correcting, if plausible, any decrease in observed scores across sessions. M3 employs a simpler approach to M1 and M2, whereby modelling the overall learning trend is done using smooth cubic functions, but it does not allow for the same flexibility in modelling individual differences. Rather, it only captures between-children differences in baseline accuracy (intercepts) and in how fast they improve (slopes), without considering more complex patterns of learning over time ^28,29^. Further details are given in the Supplementary Materials Section 5.

Population-level mean curves and Individual learning trajectories were estimated using the models M1, M2 and M3 by analysing the longitudinal data points for all participants (88 individuals), n=651, n=486, n=651 repeated measurements, respectively. The reduced number of observations considered in model M2 is due to dropping some of the repeated measurements which represented a large decrease in learning across sessions, failing to comply with our defined criterion for the monotonic model (see Supplementary Materials Section 5 for further details). To assess the robustness of our findings, we repeated our analyses using data from only the participants who completed all eight sessions (n=79), with n=632, n=471, n=632 longitudinal measurements, respectively.

We estimated associations with learning trajectories adjusted for two sets of participant characteristics: (1) child demographic and sociodemographic characteristics such as age, gender, type of school, parental education, and household income; (2) psychological traits such as verbal age, AQ50, alexithymia, learning disability, ADHD, DLD, dyslexia, and dyspraxia. First, mutually adjusted associations of demographic characteristics (i.e., association of each characteristic is adjusted for the other characteristics) with individual learning trajectories were estimated using model M1. We then estimated mutually adjusted associations of psychological traits, additionally adjusted for potential confounding from the demographic characteristics. The interaction of these characteristics with the baseline accuracy and rates of learning growth across sessions were estimated to assess the between-participants variability attributed to individual’s characteristics.

Several descriptive statistical analyses were performed to summarise emotion-specific effects over the sessions: (1) the mean accuracy for each emotion; (2) the mean number of participants’ false alarms for each emotion (i.e., the number of times an emotion was selected when this was not the correct response); (3) A-prime sensitivity scores. The A-prime sensitivity score is based on signal detection theory and is a non-parametric estimate of discriminability which captures hit and false alarm rates in a single measure of performance ^30^. The statistical summaries included the mean, 95% confidence interval of the mean, median, minimum, maximum, and range. All analyses were conducted using the R statistical software^31^.

Finally, we examined how the time taken to complete all 8 sessions impacted learning trajectories, by examining the associations using the M1 model.

## Results

### Participants

There were 88 participants in total, 59 of which were recruited from schools (across seven schools). The majority of children recruited via schools completed all the training sessions in person. Several sessions (24 sessions across 6 children) were conducted online for logistical reasons. Two children took part via the community group approach and completed sessions in person and online (3 sessions online for one child). A further 27 children took part via the social media route (211 sessions were completed online across 27 children).

Participants had a mean chronological age of 11.6 years and mean verbal age of 10.3 years. Empirical distributions of differences between chronological and verbal age are illustrated in Figure S1 in Supplementary Materials. Participants had a mean AQ50 score of 98.6 and 43% were female. Full summary statistics for participants characteristics are provided in Table 2. Data are presented for the whole sample (n=88) and separately for participants who completed all sessions (n=79), and those who completed fewer than 8 sessions (n=9).

### Evidence of Learning Effect

Accuracy scores across sessions demonstrated a consistently increasing trend (i.e., learning growth), with a mean accuracy score of 57.1 (95% CI: 54.2 to 60.0) at session S1 and 71.0 (95% CI: 68.2 to 73.9) at session S8 (see Supplementary Table S2). The distributions of accuracy scores across sessions showed a consistent improvement in learning, with reduced variability over time, see Figure 2. Individual learning trajectories are presented in Supplementary Figure S2.

**Figure 2.**
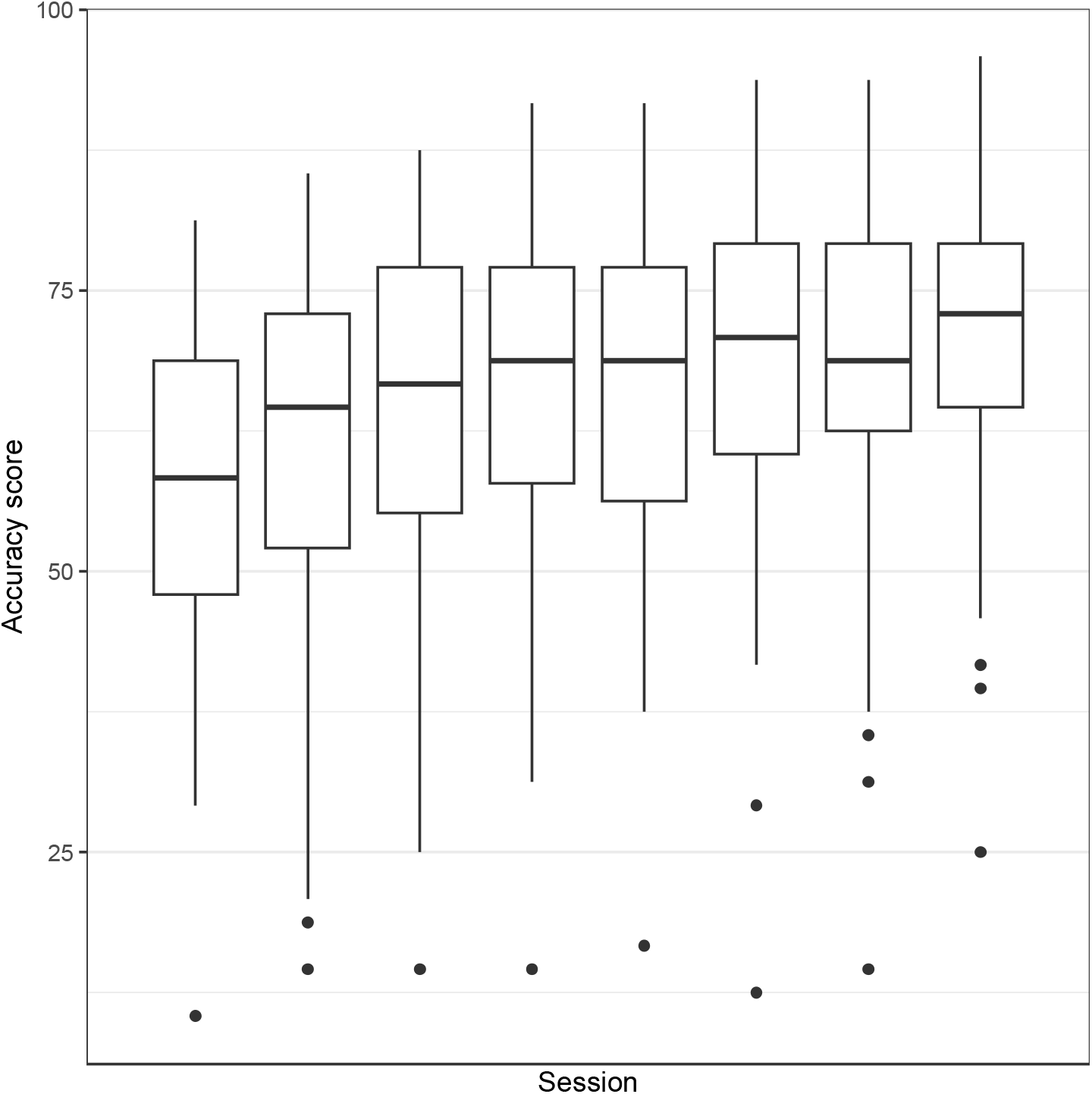
Box-plots of observed accuracy score across sessions for all participants (n = 88).

**Figure 3.**
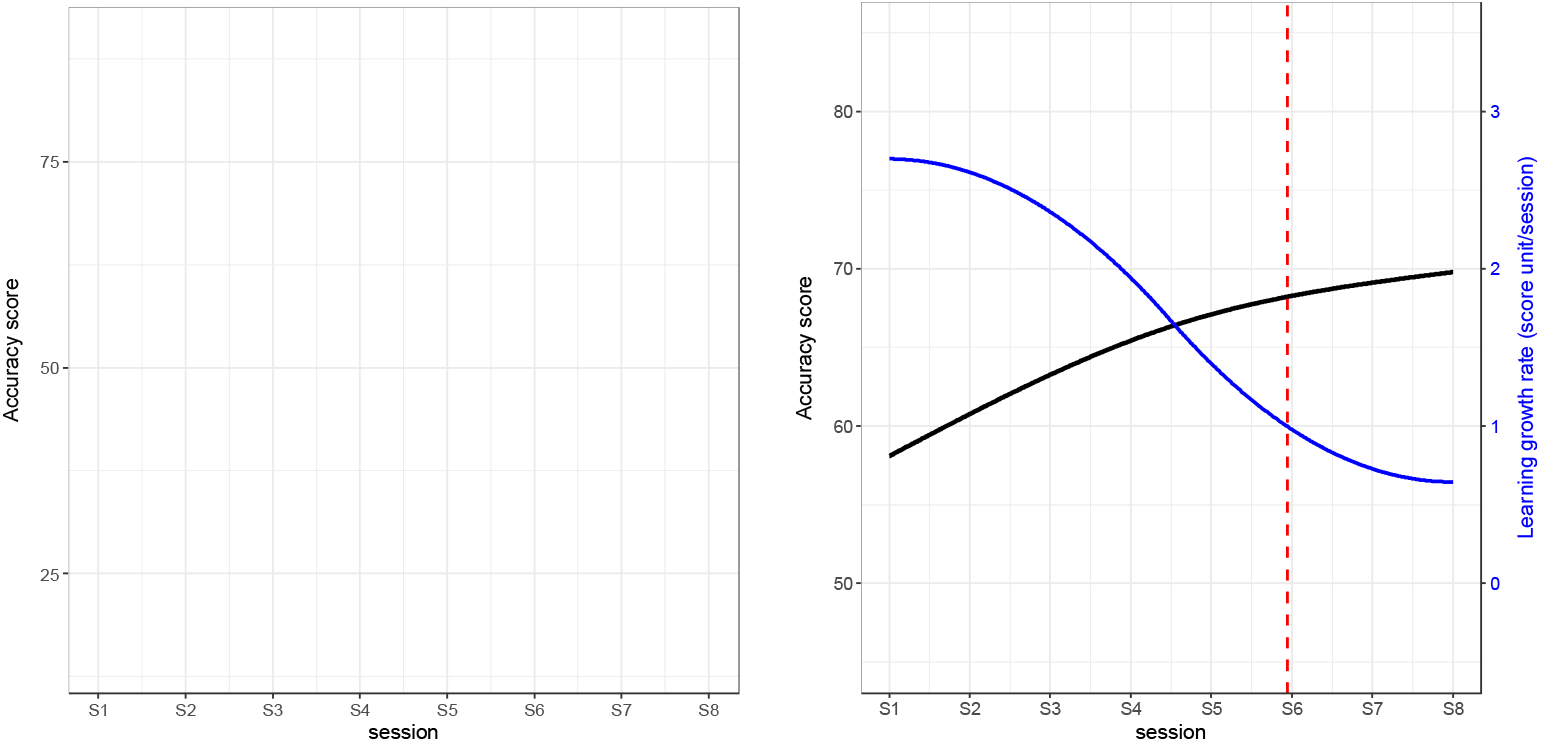
Learning growth curves estimated by model M1. Left side: Individual learning growth curves; right side: the population-mean curve (in solid black), mean learning rate (in solid blue), the optimal number of sessions (dotted red).

The population-level baseline of accuracy scores (i.e., overall mean of scores at session S1) estimated by model M1 was 58.1% (95% CI: 55.2% to 61.0%). Since we used two piece-wise smooth cubic functions, the rate of learning trajectory varies with sessions (i.e., it does not have a fixed slope across all sessions unlike a linear function). However, our model reveals that the mean rate of learning growth is larger in early sessions than in later sessions (see Table 3). Individual learning trajectories were fitted for all participants, along with the mean learning curve and rates of learning growth curve as illustrated by the black and blue lines, respectively, in Left *side:* 3. It demonstrates that the rate of learning growth decreases consistently with sessions and approaches a plateau of learning towards the latter sessions. For instance, the mean rate of learning growth was ∼ 2 units/session at S4, whereas it decreased to ∼ 0.6%/session at S8. The optimal number of sessions, defined as the number after which the mean rate of learning drops below 1 percent/session and thereby the learning plateaus, is estimated as six (see Left *side:* 3). The estimates of the model parameters of M2 and M3 and their graphical representations confirmed the results obtained from model M1, see Table 3, and Supplementary Tables S3, Figures S3 and S4.

**Table 3.** Estimates of population-level (fixed-effect) parameters for three models of accuracy scores using data from all participants (n = 88) across the eight sessions (S1 to S8)

| Parameters | M1: Main model<sup>*</sup> |  | M2: Monotonic learning model<sup>†</sup> |  | M3: Reduced model<sup>‡</sup> |  |
| --- | --- | --- | --- | --- | --- | --- |
|  | Estimate (95% CI) | p-value | Estimate (95% CI) | p-value | Estimate (95% CI) | p-value |
| Intercept<sup>§</sup> | 58.09<br>(55.17 to 61.00) | $2 \times 10^{-16}$ | 57.89<br>(55.03 to 60.75) | $2 \times 10^{-16}$ | 58.06<br>(55.10 to 61.03) | $2 \times 10^{-16}$ |
| Session<br>(1<sup>st</sup> spline: S1-S4) | 17.33<br>(12.67 to 22.00) | $2 \times 10^{-10}$ | 29.71<br>(25.24 to 34.18) | $2 \times 10^{-16}$ | 17.51<br>(13.81 to 21.20) | $2 \times 10^{-16}$ |
| Session<br>(2<sup>nd</sup> spline: S5-S8) | 7.44<br>(5.58 to 9.31) | $2 \times 10^{-11}$ | 11.25<br>(9.61 to 12.89) | $2 \times 10^{-16}$ | 7.41<br>(5.24 to 9.58) | $5 \times 10^{-10}$ |

*\*M1: mixed-effects regression spline model with natural cubic basis and one knot at session S4. The two splines are used for both fixed- and random-effect terms (the main model). †M2: same as in model M1, but the outcome of interest is monotonic non-decreasing accuracy scores, whereby an observed decrease in accuracy score between any two successive sessions is treated as missing, unless it does not exceed a certain threshold (up to -3 hits) tolerating a small performance bias, where the corresponding decreased score would be replaced by the score of the preceded session. ‡M3: mixed-effects regression spline model with natural cubic basis and one knot at session S4. The two splines are used for the fixed-effect term only, while the random-effect term represents a random slope for sessions (no splines). §the sessions are centred at session S1, then the results of intercept represent the estimated means (95% confidence interval) and p-values of accuracy scores at session S1. Abbreviations: CI - confidence interval*

Model M1 explained 89% of the learning growth variation in our study population (full sample, n=88) with residual standard deviation (SD) of 7.3 (95% CI: 6.8 to 7.8), see Supplementary Table S3. The between-participants variability in the first spline, modelling sessions 1 to 4, was the largest source of heterogeneity in individual learning trajectories with SD of 16.3 (95% CI: 11.8 to 20.9). In later sessions (5 to 8), this variability was lower with SD of 3.5 (95% CI: 1.1 to 6.3). There was also evidence of a greater rate of learning growth in earlier sessions (coefficient = 17.3 in first half of training) than later sessions (coefficient = 7.4 in second half of training), see Table 3. This is likely to reflect participants approaching a ceiling level of learning, and the point where additional training is less beneficial.

There was evidence for a negative linear correlation between individual baseline accuracy scores (at session S1) and rates of learning growth in later sessions (5 to 8) with a coefficient of -0.8 (95% CI: -1.0 to -0.3) indicating that participants with lower baseline scores are more likely to have larger rates of learning growth in late sessions. Similar findings were estimated by model M2 with coefficients of -0.5 (95% CI: -0.7 to -0.2), see Supplementary Table S3.

Moreover, there was evidence for a positive correlation between learning growth rates for earlier (1 to 4) and later (5 to 8) sessions under model M2, with coefficient of 0.9 (95% CI: 0.7 to 1.0). Model M3 did not produce evidence for a correlation between the baseline accuracy scores and growth rates (slopes) suggesting that the patterns in the data are more complex than model M3 can capture (see Supplementary Table S3).

There was no evidence for an influence of the mode of learning delivery on the estimated parameters of model M1 (see Supplementary Table S6).

When restricting our analyses to only the participants who completed all the eight sessions (n=79) for complete-case analyses, the findings from the three models were similar to the ones obtained from the whole study population in terms of population-level baseline, patterns of learning growth, and the estimated number of optimal sessions (see Supplementary Tables S4 and S5).

### Model adjustment for participant’s characteristics

#### Accuracy Score Distributions Per Participant’s Characteristics Across All Sessions

We explored the distributions of accuracy scores across sessions (learning) by different levels of investigated child characteristics: gender, age, verbal age, difference between age and verbal age, parental education, household income, school type (mainstream, SEN), AQ50, alexithymia, learning disability, ADHD, DLD, dyslexia and dyspraxia. Data for each are plotted in Supplementary Figures S5-S17. Due to very few cases of DLD, the plot for this trait is not presented due to risk of identification.

Participants with higher verbal age (> 9.25 years) had marginally higher accuracy scores across sessions. This association was more pronounced in the early sessions suggesting that participants with lower verbal ages caught up in later sessions, see Supplementary Figure S7. Participants from households with larger annual income (≥50k) had larger accuracy scores across all sessions (see Supplementary Figure S10).

#### Baseline Differences in Accuracy Score

Among the demographic and sociodemographic characteristics, there was evidence for an association of gender with accuracy score at baseline, with males having scores that were on average 7.2 percentage points (95% CI: 1.1 to 13.3; *P* = 0.03) larger than females, see Table 4. There was weak evidence that lower parental education was associated with lower baseline score (-7.7%; 95% CI: -15.2 to -0.2; *P* = 0.05) compared with higher parental education, and that higher age (>11.79 years) was associated with higher baseline score (6.6 percentage points; 95% CI: -0.3 to 13.6). There was no clear evidence of baseline differences based on other individual factors assessed.

**Table 4.** Mutually adjusted associations of participant’s characteristics with longitudinal learning trajectories, estimated by model M1* for all participants^†^

| Parameters | Estimate (95% CI) | p-value |
| --- | --- | --- |
| Intercept<sup>§</sup> | 55.03 (47.84 to 62.21) | $2 \times 10^{-16}$ |
| Session (1<sup>st</sup> spline: S1-S4) | 10.04 (-1.13 to 21.30) | 0.096 |
| Session (2<sup>nd</sup> spline: S5-S8) | 6.37 (1.65 to 11.12) | 0.009 |
| Age >11.79 years | 6.63 (-0.33 to 13.58) | 0.077 |
| Verbal age >9.25 years | 4.43 (-2.41 to 11.27) | 0.208 |
| Male | 7.22 (1.10 to 13.33) | 0.029 |
| SEN school | -3.87 (-11.42 to 3.68) | 0.336 |
| No parental educational qualifications | -3.29 (-13.27 to 6.68) | 0.535 |
| Pre-high parental educational qualifications | -7.68 (-15.15 to -0.22) | 0.046 |
| Household income ≥ £50k | -0.24 (-7.55 to 7.06) | 0.949 |
| 1<sup>st</sup> spline × Age >11.79 years | -7.95 (-18.73 to 2.81) | 0.169 |
| 2<sup>nd</sup> spline × Age >11.79 years | -6.84 (-11.36 to -2.34) | 0.004 |
| 1<sup>st</sup> spline × Male | 0.18 (-9.43 to 9.78) | 0.972 |
| 2<sup>nd</sup> spline × Male | -0.87 (-4.93 to 3.22) | 0.680 |
| 1<sup>st</sup> spline × SEN school | 1.33 (-10.30 to 12.94) | 0.829 |
| 2<sup>nd</sup> spline × SEN school | 3.86 (-1.00 to 8.71) | 0.123 |
| 1<sup>st</sup> spline × No parental educational qualifications | 5.80 (-9.62 to 21.22) | 0.481 |
| 2<sup>nd</sup> spline × No parental educational qualifications | 4.79 (-1.73 to 11.36) | 0.156 |
| 1<sup>st</sup> spline × Pre-high parental educational qualifications | 2.48 (-9.17 to 14.04) | 0.688 |
| 2<sup>nd</sup> spline × Pre-high parental educational qualifications | 2.22 (-2.69 to 7.09) | 0.378 |
| 1<sup>st</sup> spline × Household income ≥ £50k | 21.47 (10.20 to 32.77) | $7 \times 10^{-4}$ |
| 2<sup>nd</sup> spline × Household income ≥ £50k | 4.61 (-0.12 to 9.33) | 0.060 |

Rows showing data by participant's characteristic depict the association with absolute accuracy score at baseline, session S1. Rows showing data by spline depict learning growth by factor listed where 1<sup>st</sup> spline estimates learning rate over early sessions (1 to 4) and 2<sup>nd</sup> spline estimates learning rate over later sessions (5 to 8). \*M1: mixed-effects regression spline model with natural cubic basis and one knot at session S4. The two splines are used for both fixed- and random-effect terms (the main model in this study). †all participants whose data on all considered characteristics are available ( $n = 72$ ). §the intercept is centered at session S1, then the corresponding results represent the estimated means (95% confidence interval) and p-values of accuracy scores at S1 for females aged $\leq 11.79$ years attending mainstream school with higher parental education and household income $< £50k$ (i.e., for baseline levels of considered characteristics). Abbreviations: SEN - Special Educational Needs, CI - confidence interval

#### Differences in Learning Growth (Trajectory)

In addition to exploring absolute accuracy at baseline, spline results identify differences in learning growth rates between early (1 to 4) and late (5 to 8) sessions (see Table 4, 2^nd^ and 3^rd^ rows). Note that lower learning growth in later sessions indicates a potential plateauing of learning as children reach ceiling levels of learning.

Age was negatively associated with learning growth rates, with more pronounced evidence of age effects in late sessions (5 to 8); older participants (> 11.79 years) were more likely to have lower rates of learning growth in late sessions compared with younger participants as demonstrated by the estimate of the corresponding 2^nd^ spline (sessions 5 to 8) coefficient featuring the weight of this factor (age) in the underlying growth curve (coefficient = -6.8; 95% CI: -11.4 to -2.3; *P* = 0.004). There was evidence that household income was positively associated with learning growth rates. This association was more pronounced in the early sessions (1 to 4) (coefficient = 21.5 for 1^st^ spline coefficient; 95% CI: 10.2 to 32.8; *P* = 7 10^-4^; for sessions 1 to 4), see Table 4. The investigated participants’ characteristics attributed 15.7% to the between-participants variability in learning growth, see Supplementary Table S7.

There was no evidence of an association of any of the investigated autistic, alexithymia, ADHD, DLD, Dyslexia or Dyspraxia traits with learning at baseline or growth of learning, when adjusting for participants’ characteristics, see Table 5. These traits explained only ∼ 5% of between-participants variability in accuracy score and learning growth rates, see Supplementary Table S7.

**Table 5.** Mutually adjusted associations of participant’s psychological traits, additionally adjusted for participant’s characteristics*, with longitudinal learning trajectories estimated by model M1^†^ for all participants^‡^

| Parameters | Estimate (95% CI) | p-value |
| --- | --- | --- |
| Intercept<sup>§</sup> | 53.17 (42.36 to 63.98) | $2 \times 10^{-16}$ |
| Session (1<sup>st</sup> spline: S1-S4) | 7.96 (-9.03 to 24.95) | 0.360 |
| Session (2<sup>nd</sup> spline: S5-S8) | 7.10 (0.33 to 13.88) | 0.040 |
| Non-negative difference of (verbal age – age) | -0.66 (-9.12 to 7.80) | 0.880 |
| AQ50 score >100 | 3.62 (-3.91 to 11.15) | 0.350 |
| Alexithymia >24 | 0.36 (-6.99 to 7.71) | 0.920 |
| Learning disability | -3.04 (-13.11 to 7.03) | 0.550 |
| ADHD | -2.39 (-10.94 to 6.17) | 0.580 |
| Dyslexia | -3.01 (-20.32 to 14.31) | 0.730 |
| Dyspraxia | 1.99 (-10.07 to 14.05) | 0.750 |
| 1<sup>st</sup> spline × Non-negative (verbal age – age) | 4.76 (-8.50 to 18.02) | 0.480 |
| 2<sup>nd</sup> spline × Non-negative (verbal age – age) | 1.63 (-3.63 to 6.88) | 0.540 |
| 1<sup>st</sup> spline × AQ50 score >100 | 0.69 (-11.42 to 12.81) | 0.910 |
| 2<sup>nd</sup> spline × AQ50 score >100 | -2.86 (-7.71 to 1.99) | 0.250 |
| 1<sup>st</sup> spline × Alexithymia >24 | -0.41 (-12.14 to 11.32) | 0.950 |
| 2<sup>nd</sup> spline × Alexithymia >24 | 0.71 (-4.00 to 5.42) | 0.770 |
| 1<sup>st</sup> spline × Learning disability | -0.83 (-16.56 to 14.90) | 0.920 |
| 2<sup>nd</sup> spline × Learning disability | 2.00 (-4.28 to 8.28) | 0.530 |
| 1<sup>st</sup> spline × ADHD | -2.62 (-16.39 to 11.14) | 0.710 |
| 2<sup>nd</sup> spline × ADHD | 0.10 (-5.48 to 5.67) | 0.970 |
| 1<sup>st</sup> spline × Dyslexia | 14.04 (-12.62 to 40.70) | 0.300 |
| 2<sup>nd</sup> spline × Dyslexia | -8.49 (-18.91 to 1.92) | 0.110 |
| 1<sup>st</sup> spline × Dyspraxia | -8.63 (-28.31 to 11.05) | 0.390 |
| 2<sup>nd</sup> spline × Dyspraxia | 1.96 (-5.93 to 9.85) | 0.630 |

*\*the model estimates are additionally adjusted for age, gender, type of school, parental education, household income. †M1: mixed-effects regression spline model with natural cubic basis and one knot at session S4. The two splines are used for both fixed- and random-effect terms (the main model). ‡all participants whose data on all considered characteristics and traits are available (n = 72). §the intercept is centred at session S1. Abbreviations: CI - confidence interval*

### Other learning metrics

Both mean and median number of attempts (i.e., number of guesses before a correct response) and false alarms showed gradual decline with sessions, see Supplementary Tables S8 and S9, and Supplementary Figures S18 and S19. The number of attempts had a mean of 2.0 (95% CI: 1.9 to 2.1) at session 1, which was reduced to 1.6 (95% CI: 1.5 to 1.6) at session 8. The repeated measurement distributions of false alarms per emotion illustrated a consistent pattern of false alarm reduction over sessions with a clearer declining pattern for the ‘surprised’ emotion, see Supplementary Figure S19. The overall A-prime sensitivity score increased slightly across all emotions over sessions with a range of mean values from 0.8 to 0.9, see Supplementary Table S10. The gradual improvement in the A-prime scores over sessions is evident as demonstrated by the distributions per emotion, depicted in Supplementary Figure S20.

Finally, there was no evidence for an association between time to complete the training and neither baseline scores, (-0.1%; 95% CI: -0.3 to 0.02; P = 0.08), learning growth trajectories in earlier sessions (S1-S4), -0.03%; 95% CI: -0.3 to 0.19; P = 0.8), nor trajectories in later sessions (S5-S8), 0.01%; 95% CI: -0.08 to 0.1; P = 0.8) (Supplementary Table S11).

## Discussion

In this study we examined how emotion recognition accuracy scores changed over a series of 8 sessions of emotion recognition training in autistic children, or those identified as autistic by parents/carers. Our first aim was supported as we observed improvements in ER across sessions. Accuracy scores generally improved, increasing from a mean of 58 to 73 (out of 100) across the 8 sessions, representing a 25% increase in accuracy overall. The between-participant variability in scores also decreased over time suggesting that those children who started off with lower scores ‘caught up’ with their peers over time.

The second aim of this study was to examine how learning growth varied for different children. Our findings suggest that those with a higher verbal age tended to have greater overall accuracy scores and that those with a lower verbal age needed more sessions to catch up to peers with a higher verbal age. In addition, as both higher household income and higher parental education level were positively associated with increased scores/learning growth, socioeconomic factors potentially influenced performance on these tasks. Finally, female participants and younger children had lower accuracy scores at baseline. However, even when accounting for these factors there was evidence for a positive effect of the training on emotion recognition accuracy for all children.

Finally, the third aim of this work was to identify the optimal number of sessions needed for this task. We found that 6 sessions was optimal, as this was when learning tended to plateau, although this varied somewhat depending on children’s characteristics. This finding may serve as a useful guide to other studies using similar tasks as it indicates that multiple sessions are likely to be needed to achieve better outcomes up to a point, after which limited additional benefit is observed.

Our findings of improved ER after training are in line with previous studies^16,32,33^. However, of these studies many have small sample sizes and only a few included multiple training sessions. Of those studies that did include multiple sessions, they tend to align with our results reporting continued improvement, whereby a greater number of sessions resulted in greater improvements in ER. However, these studies have not examined the optimal number of sessions that are needed before accuracy scores plateau. Previous studies also did not consider in detail how learning over time relates to key participant characteristics, which our study addressed. However, we did not find associations with all of the measured participant characteristics. Given that we were working with an autistic group of schoolchildren it may be that some characteristics we examined would be more important to consider in a group of schoolchildren which also includes non-autistic individuals. For example, differences in the AQ50 may be more apparent in a group including both autistic and non-autistic individuals and therefore may have a differential impact on emotion recognition accuracy than in the current sample. In addition, there were some variables we included (e.g., other diagnoses) that had low numbers of cases. We may therefore have been underpowered to detect any differences in the effect on emotion recognition accuracy of these variables in this study.

### Strengths and Limitations

Our study has several key strengths including a large sample size for a repeated sessions study and good retention of participants resulting in complete case data for most participant. The, mixed-effects smoothing spline modelling approach is rarely used in this context, and offered a valuable and novel insight into learning growth trajectories across the sample and individuals. Thus, this informs approaches to implementing training in practice. However, there are limitations that are also important to highlight. First, we were unable to meet our initial target sample size, meaning that we may have been underpowered to detect some effects in our analyses, for example when considering different participant characteristics. Second, there was variability in the time taken to complete all 8 sessions due to variations in the spacing between them, as well as in the locations where these sessions took place. We found that there was no influence of the time taken to complete the sessions on our results. However, future studies examining the impact of this on longer term outcomes would be useful Third, our statistical models used only two splines covering different training periods, arbitrarily chosen to be from sessions 1 to 4 and from sessions 5 to 8, to capture complex non-linear relationships in the data. Although this seems a reasonable choice for our study, these two regions might have not adequately detected the entire complexity in the learning growth patterns. This is important if there were substantial variations in accuracy scores across sessions, particularly in regions far from the middle sessions. Moreover, the choice of the knot location (at the middle) to split the time into two segments may not be optimal. In extended studies with more participants and sessions, additional spline segments with an increased model flexibility should be examined. Furthermore, our definition of the optimal number of sessions as the number after which the mean rate of learning drops below 1 percent/session, i.e., plateaus, is somewhat arbitrary. Although this is not an unreasonable criterion, a different number of optimal sessions could be obtained if one considers a different criterion for optimal number of sessions.

## Conclusion

In this study we examined how ER accuracy changes over 8 sessions of an ER training task in autistic schoolchildren or those considered to be autistic. We found that learning rates increased, and variability decreased across sessions, suggesting training improved ER across most children and differences between children in ER ability was lessened. Whilst there was evidence of general improvement, some individual characteristics may influence learning. We identified that 6 sessions of the task seemed optimal. We utilised the mixed-effects smoothing spline models for statistical analyses, a modelling approach that brings novel value to this context. This research serves as a useful guide to other researchers using similar tasks in terms of aspects that might influence learning rates. Further work is needed to understand the downstream effect of improved ER on broader social and wellbeing outcomes, which has not been well studied. In addition, understanding how different participant characteristics are important for learning across all children and not just autistic children and whether the degree of learning rate is different between autistic and non-autistic children would be useful to examine in future research.

## Acknowledgements

We would like the thank all of the research participants, parents, schools, and community centres who took part in our study. Without their participation this work would not have been possible. For the purpose of open access, the author(s) has applied a Creative Commons Attribution (CC BY) licence to any Author Accepted Manuscript version arising from this submission.

## Author contributions

Conceptualization: OM, ASA, ZER; Methodology: OM, DH, ZER; Formal Analysis: OM; Investigation: OM, DH, ZER; Resources: ZER, ASA; Data Curation: DH, ZER; Writing—Original Draft: OM, ZER; Writing—Review and Editing: OM, DH, ISPV, CJ, MRM, ASA, ZER; Supervision: OM, ASA, ZER; Project Administration: ASA, ZER; Funding Acquisition: ASA, ZER.

