## Supplementary Materials for "Modelling learning trajectories of facial emotion recognition training in autistic children"

### 1. Deviations from protocol

There were some deviations from the protocol; below we list these deviations, with explanations for these changes.

- As discussed in the main paper we did not meet our target sample size of 110. However, we were able to conduct analyses with a sample size of 88.
- We originally planned to recruit from schools only but due to slow recruitment we opened recruitment up to community groups.
- The modelling approach had some differences to the planned approach:
  - We used a mixed-effects model with natural cubic splines instead of a logarithm function, as this was more appropriate given the data, where the fitted curve reflected the growth of a child’s learning across time. The learning score y_it_ was normalised to the scale ~~0-1~~ (0 – 100; instead of 0 – 1), and repeated measurements of this across all assessment sessions were used to fit a longitudinal mixed-effects model: yit = β0i + β1i_(spline1)_ . t + β2i_(spline2)_ . t + uit.

### 2. Autism spectrum quotient 50 questionnaire

The Autism spectrum quotient 50 (AQ50) questionnaire was used to assess autistic traits at baseline (1). Although there is an AQ50 Adolescent questionnaire available (2), we used the AQ50 Child for both children and adolescents in order to keep the parent’s online questionnaire consistent. Parents were asked to choose one response that best suited how each statement describes their child using one of the following responses, “Definitely Agree”, “Slightly Agree”, “Slightly Disagree” or “Definitely Disagree”. For questions: 1, 3, 8, 10, 11, 14, 15, 17, 24, 25, 27, 28, 29, 30, 31, 32, 34, 36, 37, 38, 40, 44, 47, 48, 49, 50 items are scored as follows: 0 points – “Definitely agree”, 1 point – “Slightly Agree”, 2 points – “Slightly Disagree”, 3 points – “Definitely Disagree”. For questions: 2, 4, 5, 6, 7, 9, 12, 13, 16, 18, 19, 20, 21, 22, 23, 26, 33, 35, 39, 41, 42, 43, 45, 46 items are scored in reverse, as follows: 3 points – “definitely agree”, 2 points – “Slightly Agree”, 1 point – “Slightly Disagree”, 0 points – “Definitely Disagree”. The 50 questions were:

1. S/he prefers to do things with others rather than on her/his own.
2. S/he prefers to do things the same way over and over again.
3. If s/he tries to imagine something, s/he finds it very easy to create a picture in her/his mind.
4. S/he frequently gets so strongly absorbed in one thing that s/he loses sight of other things.
5. S/he often notices small sounds when others do not.
6. S/he usually notices house numbers or similar strings of information.
7. S/he has difficulty understanding rules for polite behaviour.
8. When s/he is read a story, s/he can easily imagine what the characters might look like.
9. S/he is fascinated by dates.
10. In a social group, s/he can easily keep track of several different people’s conversations.
11. S/he finds social situations easy.
12. S/he tends to notice details that others do not.
13. S/he would rather go to a library than a birthday party.
14. S/he finds making up stories easy.
15. S/he is drawn more strongly to people than to things.
16. S/he tends to have very strong interests, which s/he gets upset about if s/he can’t pursue.
17. S/he enjoys social chit-chat.
18. When s/he talks, it isn’t always easy for others to get a word in edgeways.
19. S/he is fascinated by numbers.
20. When s/he is read a story, s/he finds it difficult to work out the characters’ intentions or feelings.
21. S/he doesn’t particularly enjoy fictional stories.
22. S/he finds it hard to make new friends.
23. S/he notices patterns in things all the time.
24. S/he would rather go to the cinema than a museum.
25. It does not upset him/her if his/her daily routine is disturbed.
26. S/he doesn’t know how to keep a conversation going with her/his peers.
27. S/he finds it easy to “read between the lines” when someone is talking to her/him.
28. S/he usually concentrates more on the whole picture, rather than the small details.
29. S/he is not very good at remembering phone numbers.
30. S/he doesn’t usually notice small changes in a situation, or a person’s appearance.
31. S/he knows how to tell if someone listening to him/her is getting bored.
32. S/he finds it easy to go back and forth between different activities.
33. When s/he talk on the phone, s/he is not sure when it’s her/his turn to speak.
34. S/he enjoys doing things spontaneously.
35. S/he is often the last to understand the point of a joke.
36. S/he finds it easy to work out what someone is thinking or feeling just by looking at their face.
37. If there is an interruption, s/he can switch back to what s/he was doing very quickly.
38. S/he is good at social chit-chat.
39. People often tell her/him that s/he keeps going on and on about the same thing.
40. When s/he was in preschool, s/he used to enjoy playing games involving pretending with other children.
41. S/he likes to collect information about categories of things (e.g. types of car, types of bird, types of train, types of plant, etc.).
42. S/he finds it difficult to imagine what it would be like to be someone else.
43. S/he likes to plan any activities s/he participates in carefully.
44. S/he enjoys social occasions.
45. S/he finds it difficult to work out people’s intentions.
46. New situations make him/her anxious.
47. S/he enjoys meeting new people.
48. S/he is good at taking care not to hurt other people’s feelings.
49. S/he is not very good at remembering people’s date of birth.
50. S/he finds it very to easy to play games with children that involve pretending.

### 3. Children’s Alexithymia Questionnaire questionnaire

We asked parents to complete the Children’s Alexithymia Questionnaire (CAM). There were 14 questions relating to behaviours in the child observed in the last three months. Parents could choose from one of four options of “Almost never”, “Sometimes”, “Often” and “Almost always”. The CAM has been previously published (3). Answers were scored as: 0 points – “Almost never”, 1 point - “Sometimes”, 2 points - “Often”, 3 points - “Almost always”. Scores were added for each item, to obtain a total score. A higher score indicates a greater number of behaviours associated with alexithymia. The questions were:

1. When asked about how he/she is feeling, instead talks about what he/she has been doing
2. Has difficulty saying he/she feels sad even though he/she looks sad
3. Talks about unimportant things/topics instead of sharing his/her feelings
4. Has long periods of little or no emotional expression, interrupted by bursts of emotional expression
5. Has difficulty saying he/she is happy even though he/she looks happy
6. Physically removes self from situations when asked to talk about feelings
7. Makes up unrelated stories when asked about his/her feelings
8. Verbal expressions of feelings do not match non-verbal expressions of feelings
9. Changes the topic of conversation when asked about his/her feelings
10. Has difficulty naming his/her positive feelings (such as joy, happiness, excitement)
11. Says “forget it” or “leave me alone” when asked about his/her feelings
12. Has trouble finding words or getting words out when talking about his/her own feelings
13. Uses few words (may just say “good” / “bad”) to describe most of his/her feelings
14. Says “I don’t know” when asked why he/she is upset

### 4. Session 8 questions around experience of ERT

Participants were asked the following questions at the end of session 8:

- Did you like the game you just played? (“No”, “A little”, “A lot”)
- Would you like to play it again soon? (“No”, “Yes”)
- Did you learn any new emotions while playing the game? (“No”, “Yes”)
- Was the game difficult? (“No”, “A little”, “A lot”)
- Did you have fun while playing the game?
- Do you need help to play the game? (“No”, “A little”, “A lot”)

### 5. Statistical models

For each participant, the learning score, $L_{i}$, at a single session, $i$ ($i=1, 2, \cdots, 8)$ was defined as the percentage correct hits in the session, and calculated as follows:

$$L_{i}=\frac{{total hits}_{i} \times100}{48} , i=1, 2, \cdots, 8.$$

Mixed-effects regression models, with natural cubic splines, two degrees of freedom, and one knot placed at the midpoint of sessions (4,5) were fitted for learning trajectories across sessions, S1 to S8. These settings implied two adjacent smooth functional curves (splines) joining each other at S4, i.e., modelling the data from sessions S1 to S4 (1^st^ spline) and from S4 to S8 (2^nd^ spline) using a 3^rd^ degree polynomial (cubic function). We employed these settings to fit three mixed-effects models with various underlying structures to examine the patterns of individual learning growth using different aspects of statistical analyses.

Firstly, our M1, ‘Main’, model included two natural cubic splines for both fixed- (population-level means) and random-effects (individual-level parameters that might differ among participants characterising variability in their longitudinal learning). This model featured the between-participants heterogeneity in learning through capturing variability in intercept (learning scores at the baseline session, S1), and individual learning growth trajectories for both spline curves, allowing for high level of growth flexibility.

Secondly, our M2, ‘Monotonic’, learning model considered a monotonic non-decreasing pattern of learning, (i.e., assuming an incremental learning process over time, whereby any decrease in observed scores across sessions could only be due to performance bias, such as disengagement, bad mode, lack of focus, etc), otherwise, it is typical to the main model. These decreases in learning scores were handled prior to fitting the M2 model such that if a decrease did not exceed a certain threshold, we replaced the score by the one achieved in its preceding measurement, otherwise it was excluded, resulting in a reduced number of observations considered in Model M2 compared to Models M1 and M3. We arbitrarily chose this threshold to be 7% (up to 3 hits) as a reasonable tolerance for such a correctable performance bias. Equation 1 demonstrates the definition of monotonic score, $L_{i}^{*}$, at session $i$ which has been analysed by the ‘Monotonic’ model.

$L_{i}^{*}=\left\{ \begin{matrix} set as missing & if L_{i}<L_{i-1}-7 \\ L_{i-1} & if L_{i-1}-7 \leq L_{i} <L_{i-1} \\ L_{i} & if i=1 or L_{i} \geq L_{i-1} \end{matrix} \right., i=1, 2, \cdots, 8$.	(1)

The structure of the fixed- and random-effects in model M2 is typically the same as in M1, but it is implemented on the longitudinal corrected (monotonic) scores, $L_{i}^{*}$, derived by Equation 1, rather than the raw learning scores, $L_{i}$.

Lastly, we fitted a mixed-effects regression, referred to as M3, ‘Reduced’, model that employed two natural cubic splines specifying fixed-effects only, while the random-effects included intercept and slope terms with no splines. In this model, the between-participants heterogeneity in learning growth was characterised by the variability in random intercept and random slope across all sessions. Table S1 summarises the structures of the three models.

In a secondary analysis, we have adjusted the parameters of these models for the mode of delivery (online or in-person) for each session, to examine the influence of session’s delivery mode on learning.

### Table S1. Summary of the statistical models structure used in the study

| **Model component** | **M1** | **M2** | **M3** |
| --- | --- | --- | --- |
| Name | Main | Monotonic | Reduced |
| Overall learning trajectory (fixed-effects) | Smooth cubic function | Smooth cubic function (using corrected learning scores) | Smooth cubic function |
| Between-participant differences (random-effects) | Smooth cubic function | Smooth cubic function (using corrected learning scores) | No smooth function |
| Within-participant differences (longitudinal growth pattern) | Captured | Captured | Reduced to simple structure of baseline and slope |
| Capability in capturing complex patterns (flexibility level) | High | High | Reduced |

### Table S2. Summary Statistics of accuracy scores at each session for all participants (n=88)

| **Session** | **N*** | **Mean (95% CI)** | **Median** | **Min** | **Max** | **Range** |
| --- | --- | --- | --- | --- | --- | --- |
| S1 | 88 | 57.1 (54.2 to 60.0) | 58.3 | 10.4 | 81.2 | 70.8 |
| S2 | 84 | 61.5 (58.2 to 64.8) | 64.6 | 14.6 | 85.4 | 70.8 |
| S3 | 83 | 64.3 (61.1 to 67.4) | 66.7 | 14.6 | 87.5 | 72.9 |
| S4 | 80 | 66.4 (63.0 to 69.7) | 68.8 | 14.6 | 91.7 | 77.1 |
| S5 | 79 | 66.6 (63.4 to 69.8) | 68.8 | 16.7 | 91.7 | 75.0 |
| S6 | 79 | 68.0 (64.8 to 71.2) | 70.8 | 12.5 | 93.8 | 81.2 |
| S7 | 79 | 69.1 (65.9 to 72.4) | 68.8 | 14.6 | 93.8 | 79.2 |
| S8 | 79 | 71.0 (68.2 to 73.9) | 72.9 | 25.0 | 95.8 | 70.8 |
| **Number of participants attended corresponding sessions. Abbreviations: CI - confidence interval* | | | | | | |

### Table S3. Standard deviations and linear correlation coefficients for individual learning trajectories (random-effects) estimated by three models using data from all participants (n=88)

| **Parameters** | **M1: Main model**^*^ | **M2: Monotonic learning model**^†^ | **M3: Reduced model**^‡^ |
| --- | --- | --- | --- |
| Intercept (SD)^#^ | 12.50 (10.33 to 14.96) | 13.26 (11.33 to 15.52) | 12.63 (10.61 to 14.98) |
| Session (SD) | N/A | N/A | 1.03 (0.62 to 1.41) |
| 1^st^ spline: session 1-4 (SD) | 16.27 (11.83 to 20.85) | 19.08 (15.69 to 22.96) | N/A |
| 2^nd^ spline: session 5-8 (SD) | 3.45 (1.06 to 6.30) | 5.54 (3.95 to 7.24) | N/A |
| Residuals (SD) | 7.32 (6.84 to 7.81) | 3.71 (3.41 to 4.06) | 7.67 (7.20 to 8.19) |
| Intercept & session (Cor.) | N/A | N/A | -0.29 (-0.56 to 0.09) |
| Intercept & 1^st^ spline (Cor.) | -0.28 (-0.53 to 0.04) | -0.46 (-0.63 to -0.26) | N/A |
| Intercept & 2^nd^ spline (Cor.) | -0.82 (-1.00 to -0.25) | -0.51 (-0.72 to -0.24) | N/A |
| 1^st^ spline & 2^nd^ spline (Cor.) | 0.62 (-0.10 to 0.97) | 0.89 (0.67 to 0.97) | N/A |
| Explained variation^§^ | 88.9% | 97.6% | 73.2% |
| N^¶^ | 651 | 486 | 651 |
| *All estimates are presented with their 95% confidence intervals shown within brackets.*mixed-effects regression spline model with natural cubic basis and one knot at session S4. The two splines are used for both fixed- and random-effect terms (the ‘Main’ model). †same as in the main model, but the outcome of interest is monotonic non-decreasing patterns of learning scores, whereby an observed decrease in learning score between any two successive sessions is treated as missing, unless it does not exceed a certain threshold (up to -3 hits) tolerating a small performance bias, where the corresponding decreased score would be replaced by the score of the preceded session. ‡mixed-effects regression spline model with natural cubic basis and one knot at session S4. The two splines are used for the fixed-effect term only, while the random-effect term represents a random slope for sessions (no splines). #: the intercept is centred at session S1. §calculated as the total variance (SD^2^) of random-effects in a model divided by the overall total variance, additionally including the variance of residuals. ¶the number of longitudinal observations considered in the corresponding models, where the reduced number of observations considered in model M2 is due to dropping some of the repeated measurements which represented a large decrease in learning across sessions, failing to comply with our defined critiron of monotonic model, see equation (1). Abbreviations: CI - confidence interval, N/A - not available, SD - standard deviation, Cor. = correlation coefficients.* | | | |

### Table S4. Overall means (fixed-effects) of learning trajectories estimated by model M1 using data from all participants (n=88) adjusting for the mode of delivery (in-person/online)

| **Parameters** | **M1: Main model**^*^ | |
| --- | --- | --- |
|  | **Estimate (95% CI)** | **p-value** |
| Intercept^§^ | 58.05 (54.97 to 61.12) | 2$\times$10^-16^ |
| Session (1^st^ spline: S1-S4) | 17.32 (12.60 to 22.06) | 2$\times$10^-10^ |
| Session (2^nd^ spline: S5-S8) | 7.44 (5.57 to 9.32) | 2$\times$10^-11^ |
| Online session | 0.12 (-3.37 to 3.59) | 0.943 |

**mixed-effects regression spline model with natural cubic basis and one knot at session S4. The two splines are used for both fixed- and random-effect terms (the main model). §the intercept is centred at session S1, then the corresponding results represent the estimated means (95% confidence interval) and p-values of learning scores at S1 for online sessions. Abbreviations: CI - confidence interval.*

### Table S5. Standard deviations and linear correlation coefficients for individual learning trajectories (random-effects) estimated by three models using complete-case data (n=79)

| **Parameters** | **M1: Main model**^*^ | **M2: Monotonic learning model**^†^ | **M3: Reduced model**^‡^ |
| --- | --- | --- | --- |
| Intercept (SD)^#^ | 12.56 (10.28 to 15.17) | 13.27 (11.24 to 15.67) | 12.58 (10.47 to 15.04) |
| Session (SD) | N/A | N/A | 1.04 (0.62 to 1.42) |
| 1^st^ spline: S1-S4 (SD) | 16.51 (12.02 to 21.21) | 18.99 (15.58 to 22.90) | N/A |
| 2^nd^ spline: S5-S8 (SD) | 3.45 (1.01 to 6.29) | 5.55 (3.96 to 7.25) | N/A |
| Residuals (SD) | 7.31 (6.83 to 7.81) | 3.70 (3.40 to 4.05) | 7.67 (7.19 to 8.17) |
| Intercept & session (Cor.) | N/A | N/A | -0.31 (-0.58 to 0.06) |
| Intercept & 1^st^ spline (Cor.) | -0.32 (-0.56 to 0.00) | -0.48 (-0.64 to -0.27) | N/A |
| Intercept & 2^nd^ spline (Cor.) | -0.81 (-0.99 to -0.24) | -0.52 (-0.73 to -0.24) | N/A |
| 1^st^ spline & 2^nd^ spline (Cor.) | 0.64 (-0.07 to 1.00) | 0.89 (0.67 to 0.97) | N/A |
| N^§^ | 632 | 471 | 632 |
| *All estimates are presented with their 95% confidence intervals shown within brackets. *mixed-effects regression spline model with natural cubic basis and one knot at session S4. The two splines are used for both fixed- and random-effect terms (the ‘Main’ model). †same as in the main model, but the outcome of interest is monotonic non-decreasing patterns of learning scores, whereby an observed decrease in learning score between any two successive sessions is treated as missing, unless it does not exceed a certain threshold (up to -3 hits) tolerating a small performance bias, where the corresponding decreased score would be replaced by the score of the preceded session. ‡mixed-effects regression spline model with natural cubic basis and one knot at session S4. The two splines are used for the fixed-effect term only, while the random-effect term represents a random slope for sessions (no splines). #the intercept is centred at session S1. §the number of longitudinal observations considered in the corresponding models, where the reduced number of observations considered in model M2 is due to dropping some of the repeated measurements which represented a large decrease in learning across sessions, failing to comply with our defined critiron of monotonic model, see equation (1). Abbreviations: CI - confidence interval, N/A - not available, SD - standard deviation, Cor. = correlation coefficients.* | | | |

### Table S6. Overall means (fixed-effects) of learning trajectories estimated by three models using complete-case data (n=79)

| **Parameters** | **M1: Main model**^*^ | | **M2: Monotonic learning model**^†^ | | **M3: Reduced model**^‡^ | |
| --- | --- | --- | --- | --- | --- | --- |
|  | **Estimate (95% CI)** | **p-value** | **Estimate (95% CI)** | **p-value** | **Estimate (95% CI)** | **p-value** |
| Baseline accuracy (intercept)^§^ | 58.38 (55.30 to 61.46) | 2$\times$10^-16^ | 58.14 (55.13 to 61.16) | 2$\times$10^-16^ | 58.38 (55.27 to 61.49) | 2$\times$10^-16^ |
| Session  (1^st^ spline: S1-S4) | 17.86 (13.06 to 22.65) | 2$\times$10^-10^ | 29.84 (25.28 to 34.40) | 2$\times$10^-16^ | 17.86 (14.09 to 21.62) | 2$\times$10^-16^ |
| Session  (2^nd^ spline: S5-S8) | 7.34 (5.47 to 9.21) | 4$\times$10^-11^ | 11.24 (9.59 to 12.90) | 2$\times$10^-16^ | 7.34 (5.16 to 9.52) | 8$\times$10^-10^ |
| **mixed-effects regression spline model with natural cubic basis and one knot at session S4. The two splines are used for both fixed- and random-effect terms (the main model). †same as in the main model, but the outcome of interest is monotonic non-decreasing patterns of learning scores, whereby an observed decrease in learning score between any two successive sessions is treated as missing, unless it does not exceed a certain threshold (up to -3 hits) tolerating a small performance bias, where the corresponding decreased score would be replaced by the score of the preceded session. ‡:mixed-effects regression spline model with natural cubic basis and one knot at session S4. The two splines are used for the fixed-effect term only, while the random-effect term represents a random slope for sessions (no splines). §the intercept is centred at session S1, then the corresponding results represent the estimated means (95% confidence interval) and p-values of learning scores at session S1. Abbreviations: CI - confidence interval.* | | | | | | |

### Table S7. Standard deviations for individual learning trajectories (random-effects) estimates by model M1 adjusted for different sets of characteristics

| **Parameters** | **M1: Main model**^*^ | **Model M1 adjusted for characteristics**^†^ | **Model M1 adjusted for psychological traits**^‡^ |
| --- | --- | --- | --- |
| Intercept (SD)^#^ | 12.50 (10.33 to 14.96) | 11.95 (9.42 to 13.91) | 12.44 (9.17 to 13.40) |
| 1^st^ spline: S1-S4 (SD) | 16.27 (11.83 to 20.85) | 14.67 (8.35 to 18.40) | 15.73 (7.52 to 17.65) |
| 2^nd^ spline: S5-S8 (SD) | 3.45 (1.06 to 6.30) | 2.60 (0.36 to 5.11) | 2.60 (0.18 to 4.60) |
| Residuals (SD) | 7.32 (6.84 to 7.81) | 7.35 (6.80 to 7.82) | 7.37 (6.75 to 7.75) |
| Between-participants variability explained by investigated factors (%) | N/A | 15.7% | 5.5% |
| N^§^ | 651 | 530 | 530 |
| *Estimates are presented with their 95% confidence intervals shown within brackets.*mixed-effecs regression spline model with natural cubic basis and one knot at session S4. The two splines are used for both fixed- and random-effect terms (the ‘Main’ model). †characteristics included age, gender, type of school, parental education, household income. ‡psychological traits included verbal age, AQ50, alexithymia, learning disability, ADHD, developmental language disorder, dyslexia, dyspraxia. #: the intercept is centred at session S1. §number of longitudinal observations considered in the corresponding models. The reduced number of observations in the adjusted models is due to the inclusion of covariates that are not measured for all participants. Abbreviations: CI - confidence interval, N/A = not available, SD = standard deviation.* | | | |

### Table S8. Summary statistics of number of attempts at each session for all participants (n=88)

| **Session** | **N*** | **Mean (95% CI)** | **Median** | **Min** | **Max** | **Range** |
| --- | --- | --- | --- | --- | --- | --- |
| S1 | 88 | 2.0 (1.9 to 2.1) | 1.9 | 1.4 | 3.8 | 2.4 |
| S2 | 84 | 1.9 (1.8 to 2.0) | 1.8 | 1.3 | 3.5 | 2.3 |
| S3 | 83 | 1.8 (1.7 to 1.8) | 1.6 | 1.2 | 3.5 | 2.2 |
| S4 | 80 | 1.7 (1.6 to 1.8) | 1.6 | 1.1 | 3.4 | 2.3 |
| S5 | 79 | 1.7 (1.6 to 1.8) | 1.6 | 1.1 | 3.5 | 2.4 |
| S6 | 79 | 1.7 (1.6 to 1.8) | 1.6 | 1.2 | 3.5 | 2.4 |
| S7 | 79 | 1.7 (1.6 to 1.7) | 1.6 | 1.1 | 3.5 | 2.4 |
| S8 | 79 | 1.6 (1.5 to 1.6) | 1.5 | 1.1 | 2.9 | 1.8 |
| **Number of participants attended corresponding sessions. Abbreviations: CI - confidence interval.* | | | | | | |

### Table S9. Summary statistics of mean of false alarms at each session for all participants (n=88)

| **Session** | **N*** | **Mean (95% CI)** | **Median** | **Min** | **Max** | **Range** |
| --- | --- | --- | --- | --- | --- | --- |
| S1 | 88 | 3.4 (3.2 to 3.7) | 3.3 | 1.5 | 7.2 | 5.7 |
| S2 | 84 | 3.1 (2.8 to 3.3) | 2.8 | 1.2 | 6.8 | 5.7 |
| S3 | 83 | 2.8 (2.6 to 3.1) | 2.7 | 1.0 | 6.8 | 5.8 |
| S4 | 80 | 2.7 (2.4 to 3.0) | 2.5 | 0.7 | 6.8 | 6.2 |
| S5 | 79 | 2.7 (2.4 to 2.9) | 2.5 | 0.7 | 6.7 | 6.0 |
| S6 | 79 | 2.6 (2.3 to 2.8) | 2.3 | 0.5 | 7.0 | 6.5 |
| S7 | 79 | 2.5 (2.2 to 2.7) | 2.5 | 0.5 | 6.8 | 6.3 |
| S8 | 79 | 2.3 (2.1 to 2.5) | 2.2 | 0.3 | 6.0 | 5.7 |
| **Number of participants attended corresponding sessions. Abbreviations: CI - confidence interval.* | | | | | | |

### Table S10. Summary statistics of A-prime sensitivity score at each session for all participants (n=88)

| **Session** | **N*** | **Mean (95% CI)** | **Median** | **Min** | **Max** | **Range** |
| --- | --- | --- | --- | --- | --- | --- |
| S1 | 88 | 0.8 (0.8 to 0.8) | 0.8 | 0.3 | 0.9 | 0.6 |
| S2 | 84 | 0.8 (0.8 to 0.9) | 0.9 | 0.4 | 1.0 | 0.6 |
| S3 | 83 | 0.9 (0.8 to 0.9) | 0.9 | 0.4 | 1.0 | 0.5 |
| S4 | 80 | 0.9 (0.9 to 0.9) | 0.9 | 0.3 | 1.0 | 0.6 |
| S5 | 79 | 0.9 (0.9 to 0.9) | 0.9 | 0.5 | 1.0 | 0.5 |
| S6 | 79 | 0.9 (0.9 to 0.9) | 0.9 | 0.3 | 1.0 | 0.7 |
| S7 | 79 | 0.9 (0.9 to 0.9) | 0.9 | 0.4 | 1.0 | 0.6 |
| S8 | 79 | 0.8 (0.8 to 0.8) | 0.8 | 0.3 | 0.9 | 0.6 |
| **Number of participants attended corresponding sessions. Abbreviations: CI - confidence interval.* | | | | | | |

### Table S11. Association of training duration with longitudinal learning trajectories, estimated by model M1* from participants who completed all sessions (n=79)

| **Parameters** | **M1: Main model**^*^ | |
| --- | --- | --- |
|  | **Estimate (95% CI)** | **p-value** |
| Intercept^§^ | 63.51 (56.98 to 70.03) | 2$\times$10^-16^ |
| Session (1^st^ spline: S1-S4) | 19.06 (8.70 to 29.41) | 3$\times$10^-4^ |
| Session (2^nd^ spline: S5-S8) | 6.93 (2.88 to 10.97) | 8$\times$10^-4^ |
| Training duration | -0.12 (-0.26 to 0.02) | 0.081 |
| 1^st^ spline × Training duration | -0.03 (-0.25 to 0.19) | 0.798 |
| 2^nd^ spline × Training duration | 0.01 (-0.08 to 0.10) | 0.821 |

**mixed-effects regression spline model with natural cubic basis and one knot at session S4. The two splines are used for both fixed- and random-effect terms (the main model). Abbreviations: CI - confidence interval.*

### Figure S1. Distribution of the difference between verbal age and age for all participants (n= 88)

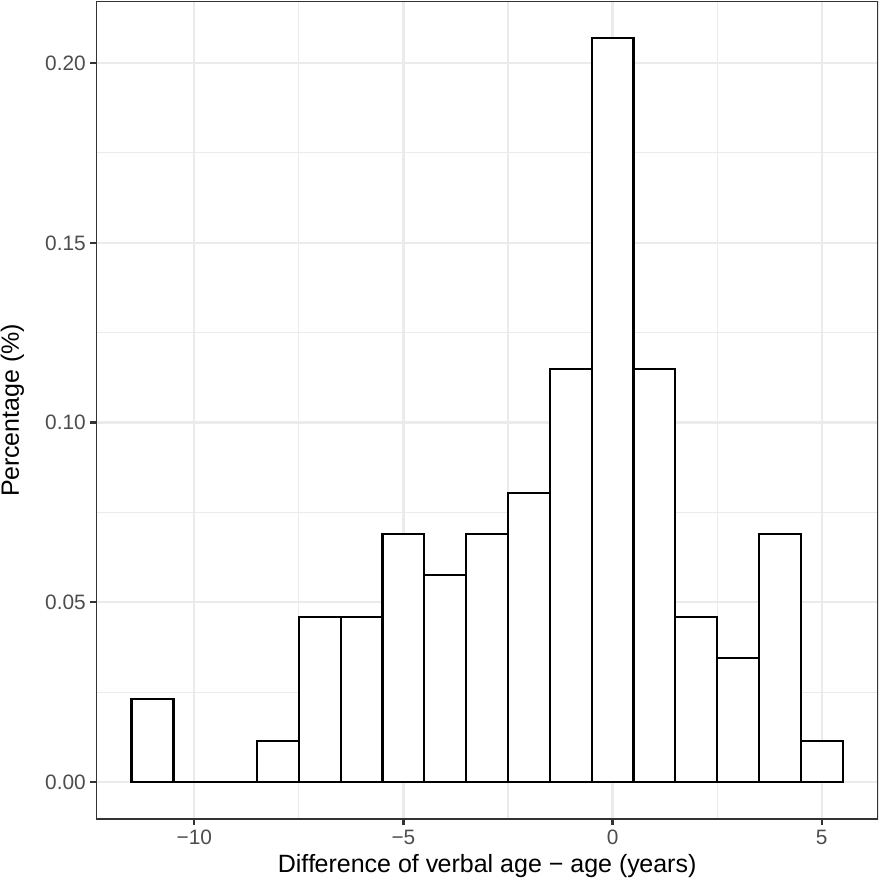

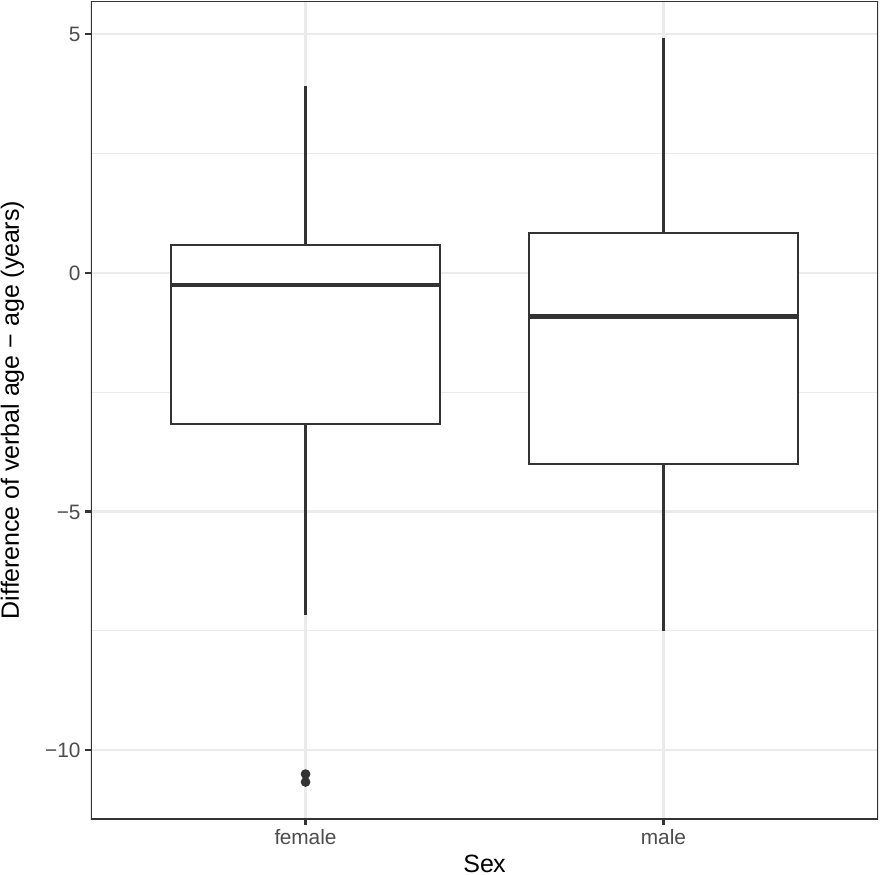

*Histogram of the difference between verbal age and age in years (left panel), and the box-plots of the difference for females and males respectively (right panel) for all participants (n=88).*

### Figure S2. Longitudinal plot of observed learning trajectories for all participants (n=88)

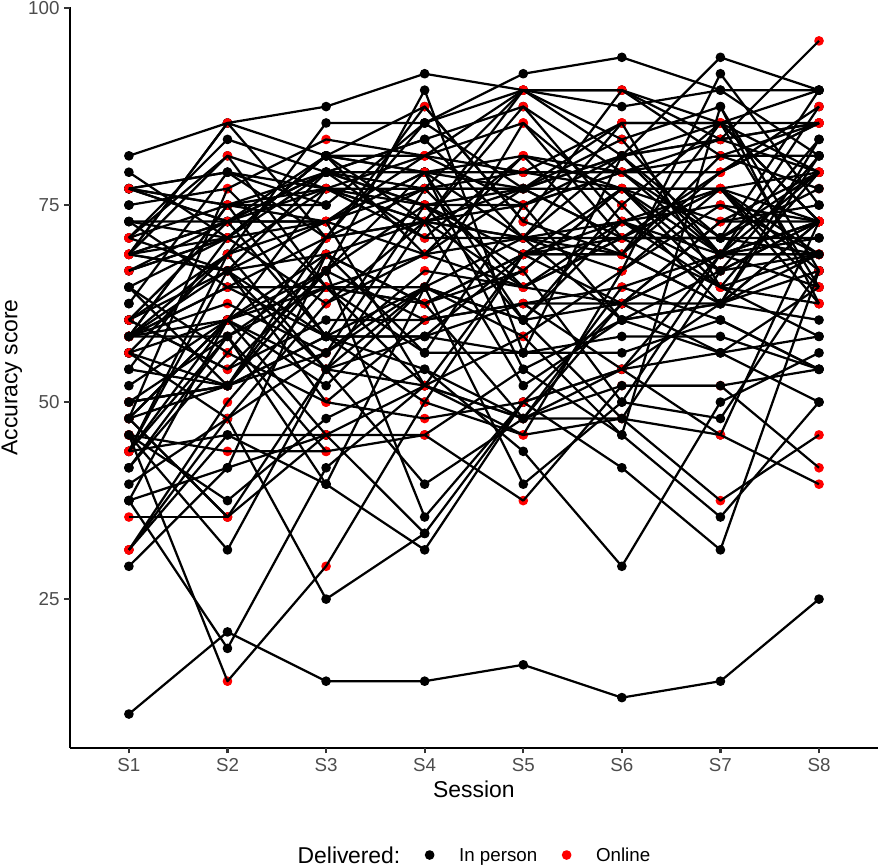

*Longitudinal plot of accuracy scores measured across the eight sessions. The mode of delivery of each session is represented by the black and red dots for ‘In person’ and ‘Online’ sessions, respectively. There was one outlier with consistently low accuracy scores. Despite a limited learning growth, this individual did show slight improvement in ER from the first to last session.*

### Figure S3. Estimated learning growth curves by the ‘Monotonic’ model

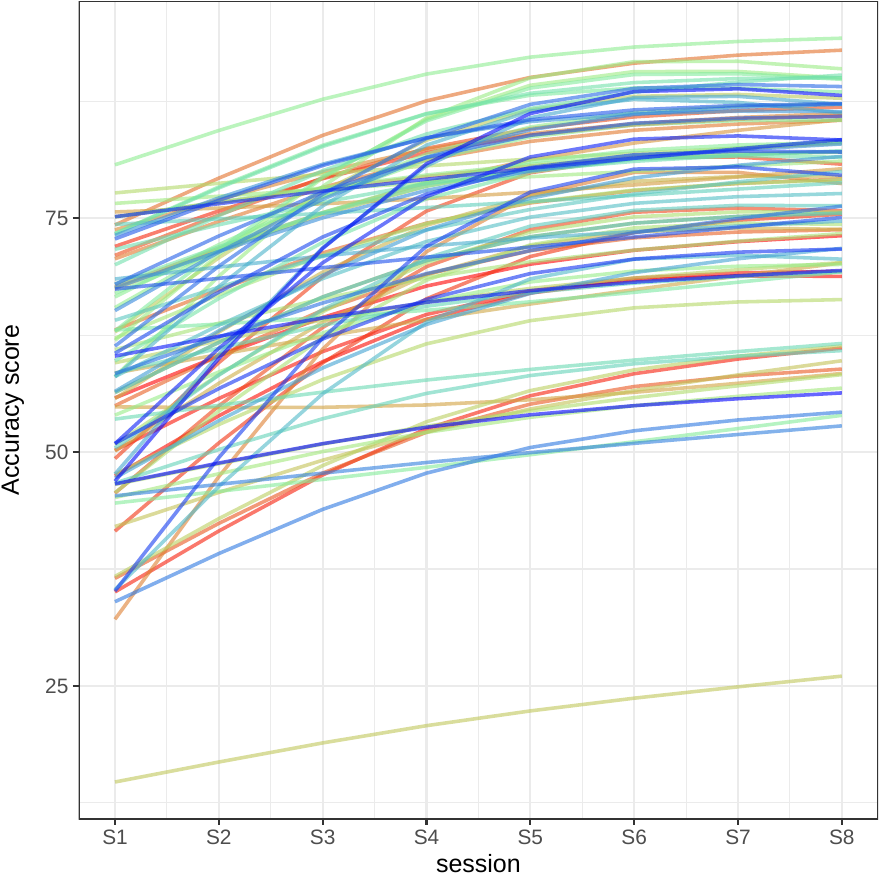

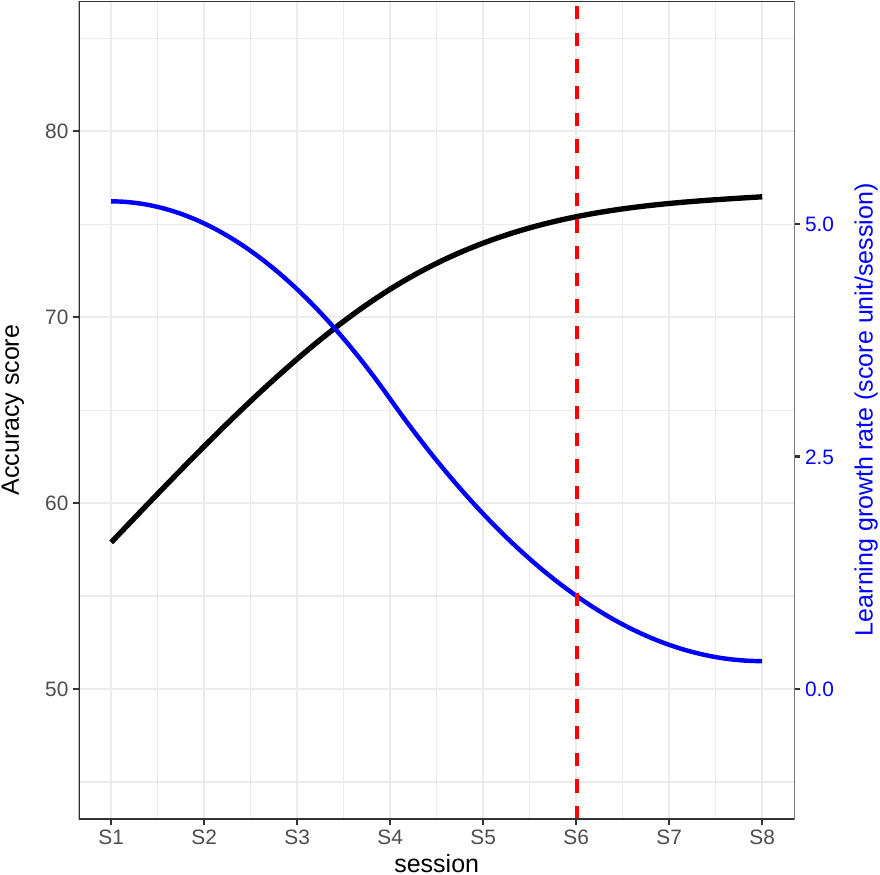

*Individual learning growth curves (left side) for all participants (n=88) with 486 measurements from eight sessions S1 to S8, and the mean curve (right side: in solid black), fitted by a mixed-effects regression model with natural cubic splines for monotonic non-decreasing accuracy scores (the ‘Monotonic’ learning model). The mean learning velocity (right side: in solid blue) is illustrated, with the vertical line (right side: in dotted red) indicating the optimal number of learning sessions (6 sessions), defined as the first session after which learning velocity falls below 1 percent/session.*

### Figure S4. Estimated learning growth curves by the ‘Reduced’ model

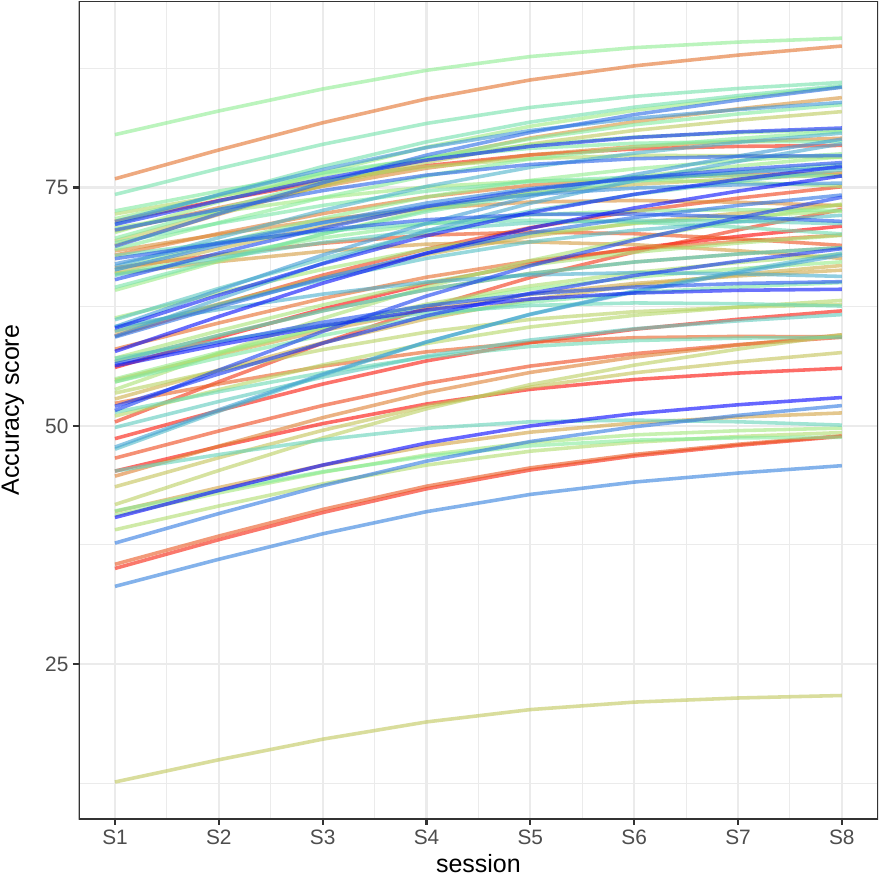

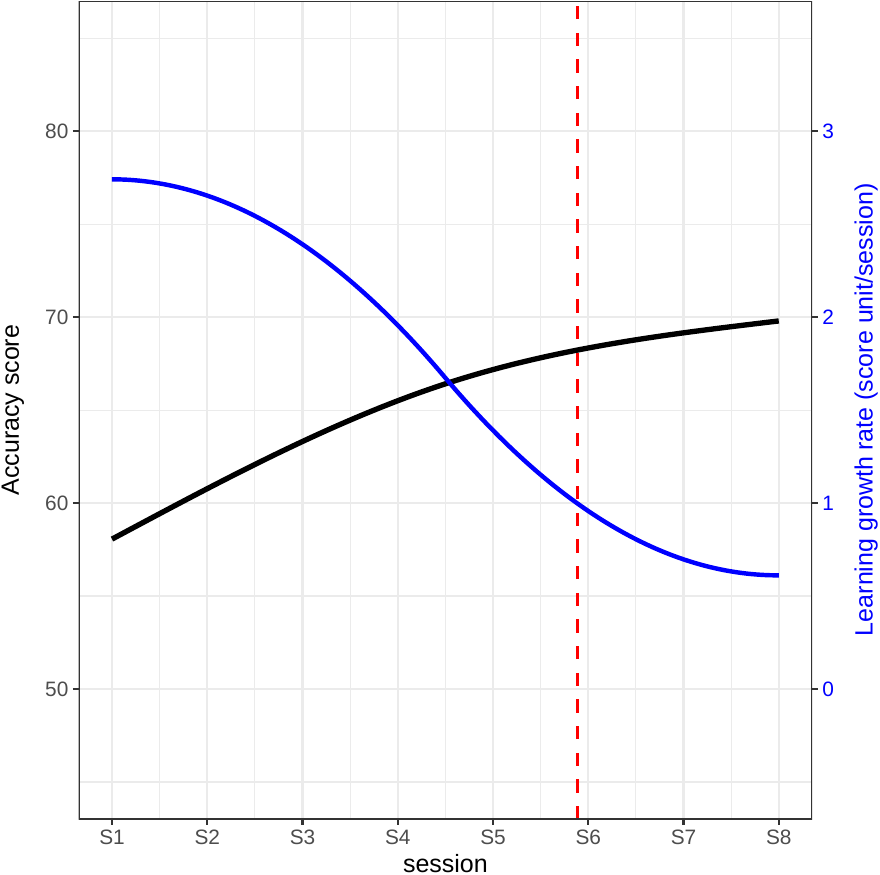

*Individual learning growth curves (left side) for all participants (n=88) with 651 measurements from eight sessions S1 to S8, and the mean curve (right side: in solid black), fitted by a mixed-effects regression model with random slope and fixed-effects of natural cubic splines (the ‘Reduced’ model). The mean learning velocity (right side: in solid blue) is illustrated, with the vertical line (right side: in dotted red) indicating the optimal number of learning sessions (6 sessions), defined as the first session after which learning velocity falls below 1 percent/session.*

### Figure S5. Scatterplot matrix of accuracy score across all pairs of sessions by gender

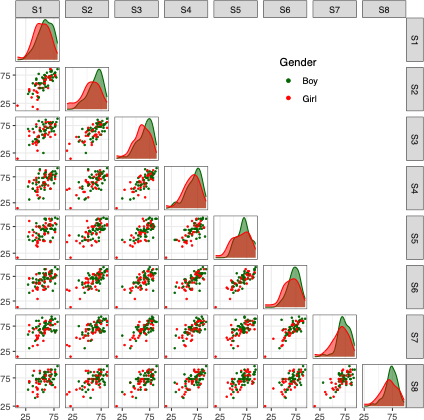

*Scatterplot matrix of accuracy score by gender for all participannts (n=88). The density plots of accuracy scores at corresponding sessions are depicted on the diagonal for each level of the distinguishing factor.*

### Figure S6. Scatterplot matrix of accuracy score across all pairs of sessions by age

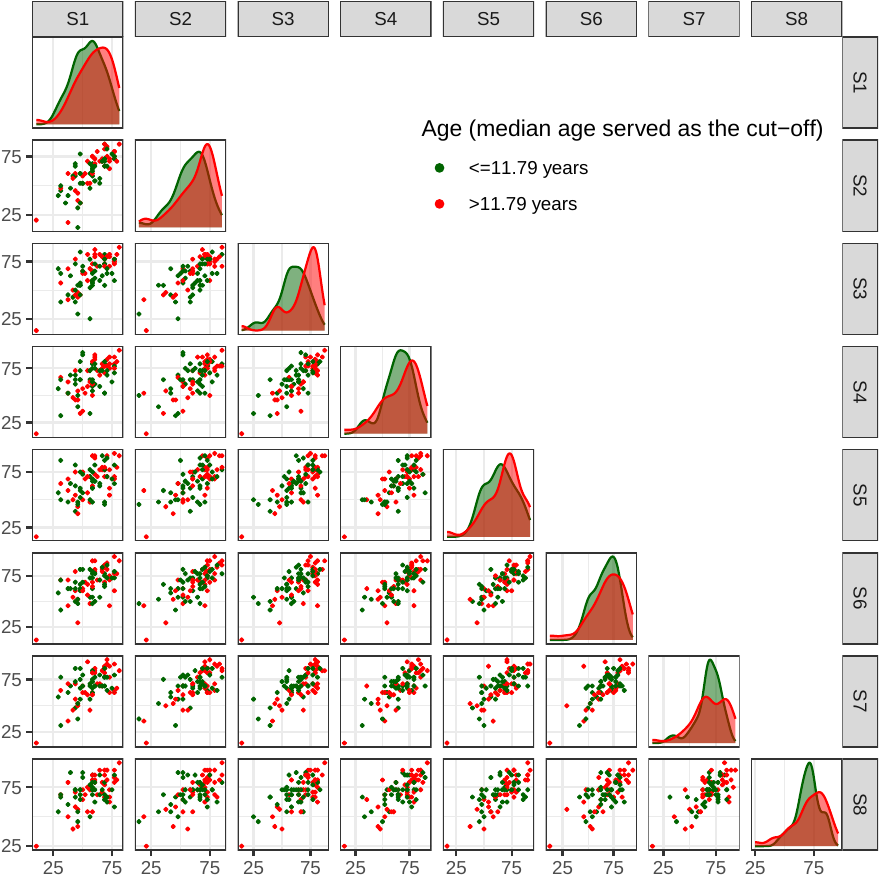

*Scatterplot matrix of accuracy score by age for all participants (n=88). The density plots of accuracy scores at corresponding sessions are depicted on the diagonal for each level of the distinguishing factor.*

### Figure S7. Scatterplot matrix of accuracy score across all pairs of sessions by verbal age

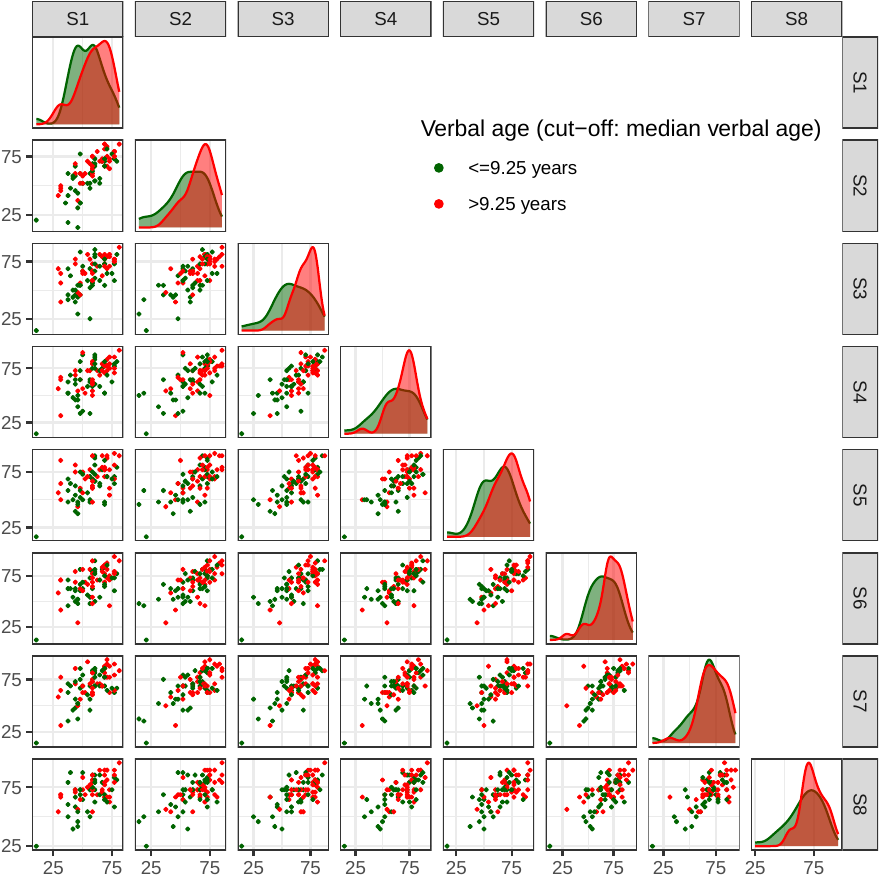

*Scatterplot matrix of accuracy score by verbal age for all participants (n=88). The density plots of accuracy scores at corresponding sessions are depicted on the diagonal for each level of the distinguishing factor.*

### Figure S8. Scatterplot matrix of accuracy score across all pairs of sessions by the difference between verbal age and age

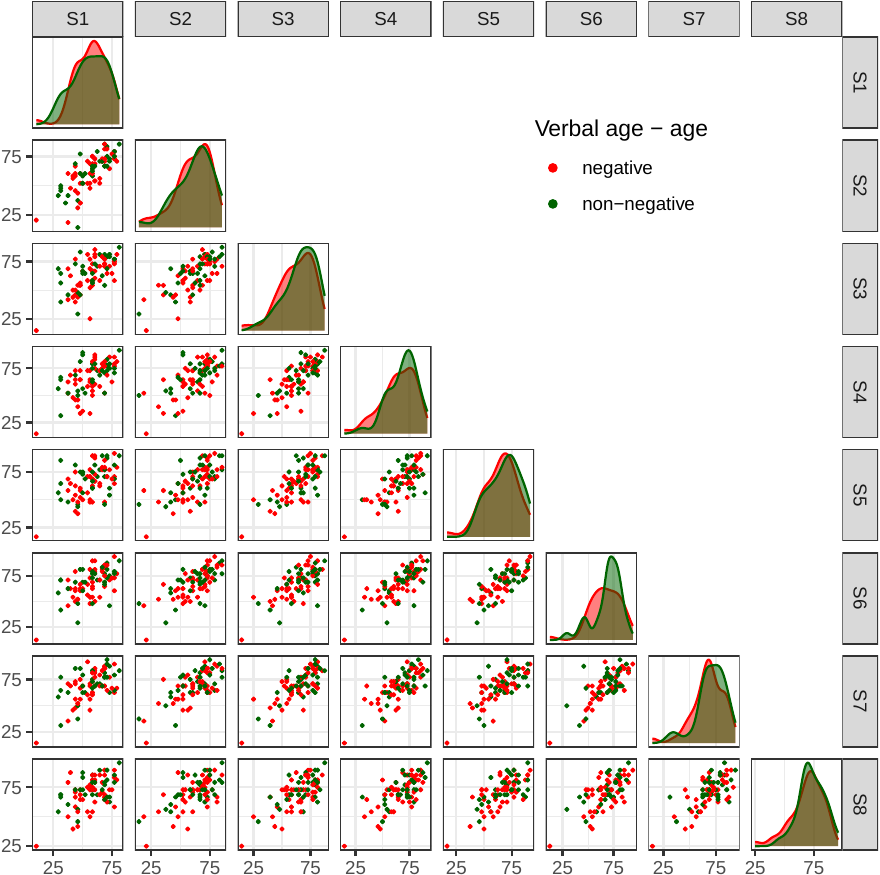

*Scatterplot matrix of accuracy score by the difference between verbal age and age for all participants (n=88). The density plots of accuracy scores at corresponding sessions are depicted on the diagonal for each level of the distinguishing factor.*

### Figure S9. Scatterplot matrix of accuracy score across all pairs of sessions by parental education

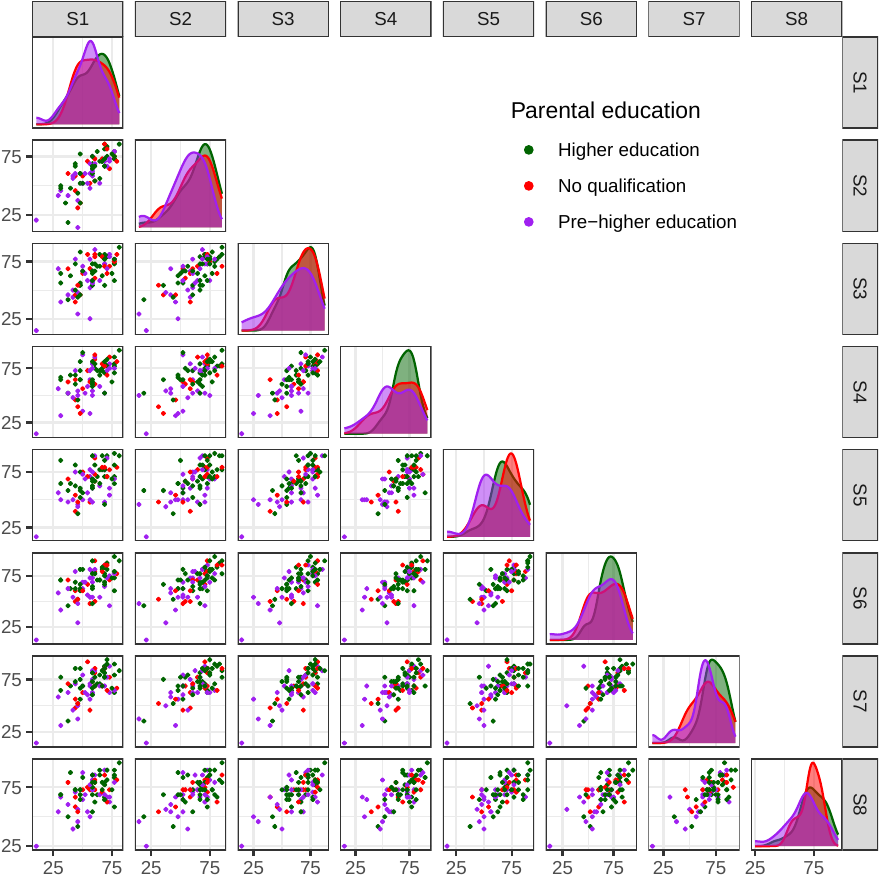

*Scatterplot matrix of accuracy score by parental education for all participants (n=88). The density plots of accuracy scores at corresponding sessions are depicted on the diagonal for each level of the distinguishing factor.*

### Figure S10. Scatterplot matrix of accuracy score across all pairs of sessions by household income

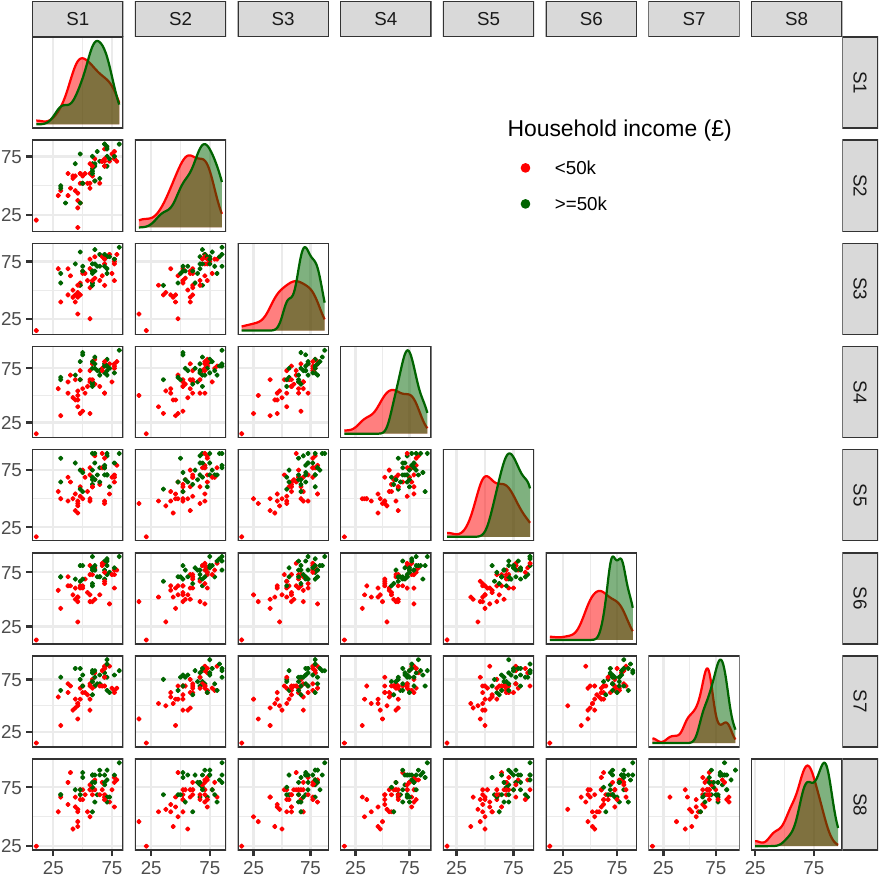

*Scatterplot matrix of accuracy score by household income for all participants (n=88). The density plots of accuracy scores at corresponding sessions are depicted on the diagonal for each level of the distinguishing factor.*

### Figure S11. Scatterplot matrix of accuracy score across all pairs of sessions by school type

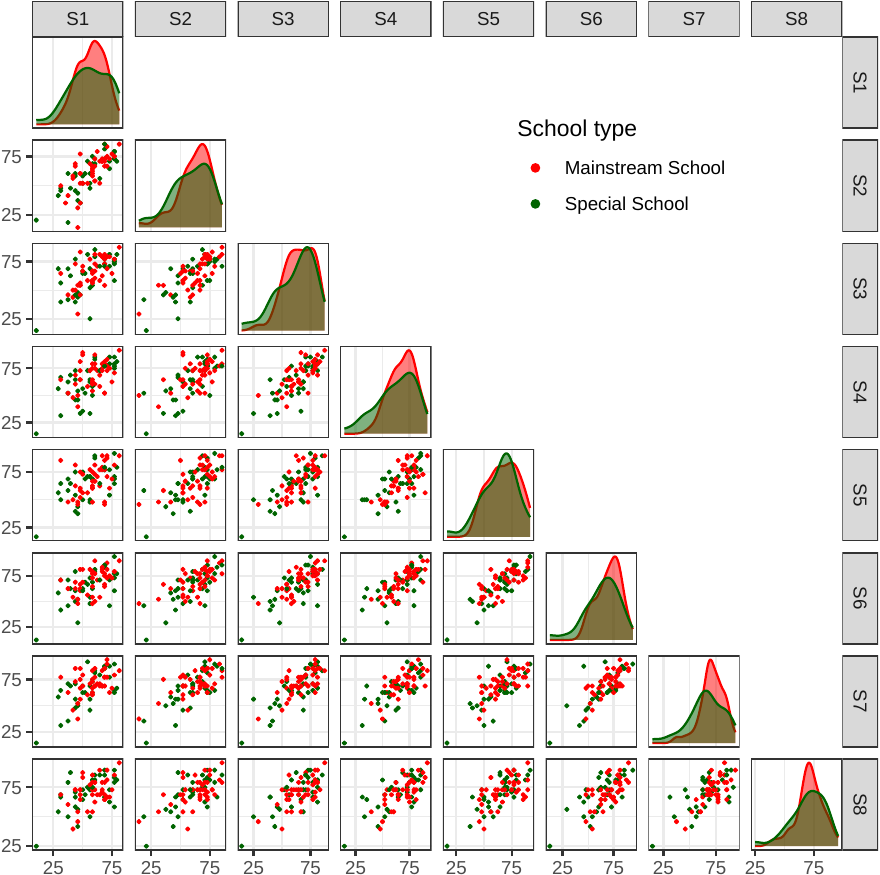

*Scatterplot matrix of accuracy score by school type for all participants (n=88). The density plots of accuracy scores at corresponding sessions are depicted on the diagonal for each level of the distinguishing factor.*

### Figure S12. Scatterplot matrix of accuracy score across all pairs of sessions by AQ50 score

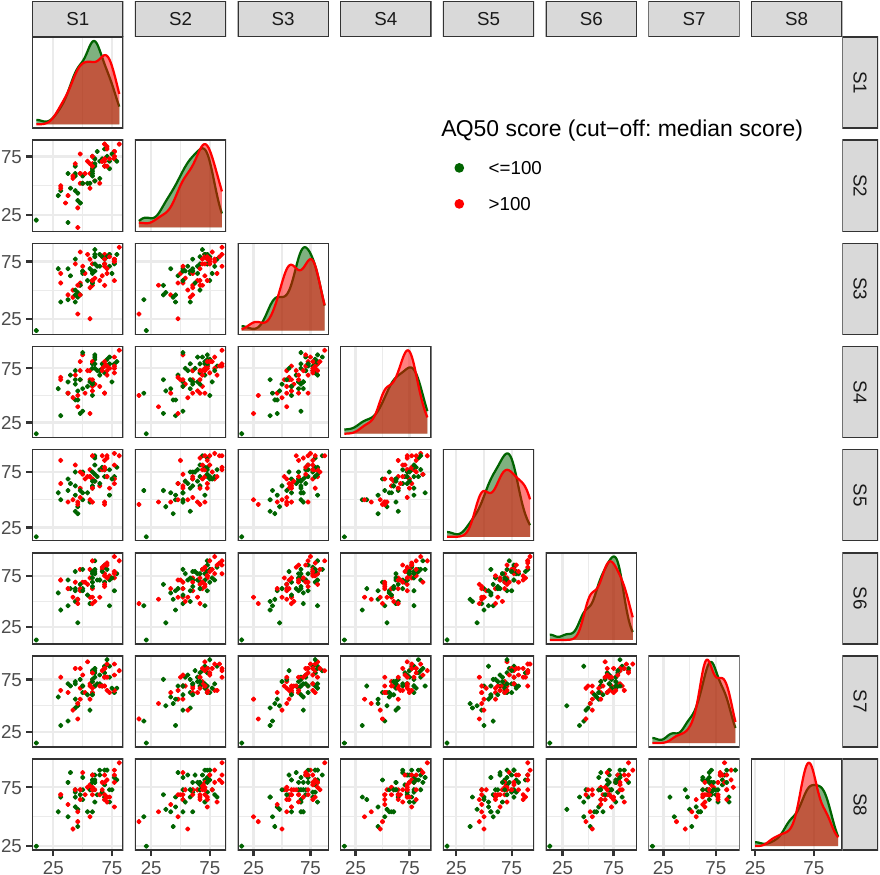

*Scatterplot matrix of accuracy score by AQ50 score for all participants (n=88). The density plots of accuracy scores at corresponding sessions are depicted on the diagonal for each level of the distinguishing factor.*

### Figure S13. Scatterplot matrix of accuracy score across all pairs of sessions by alexithymia score

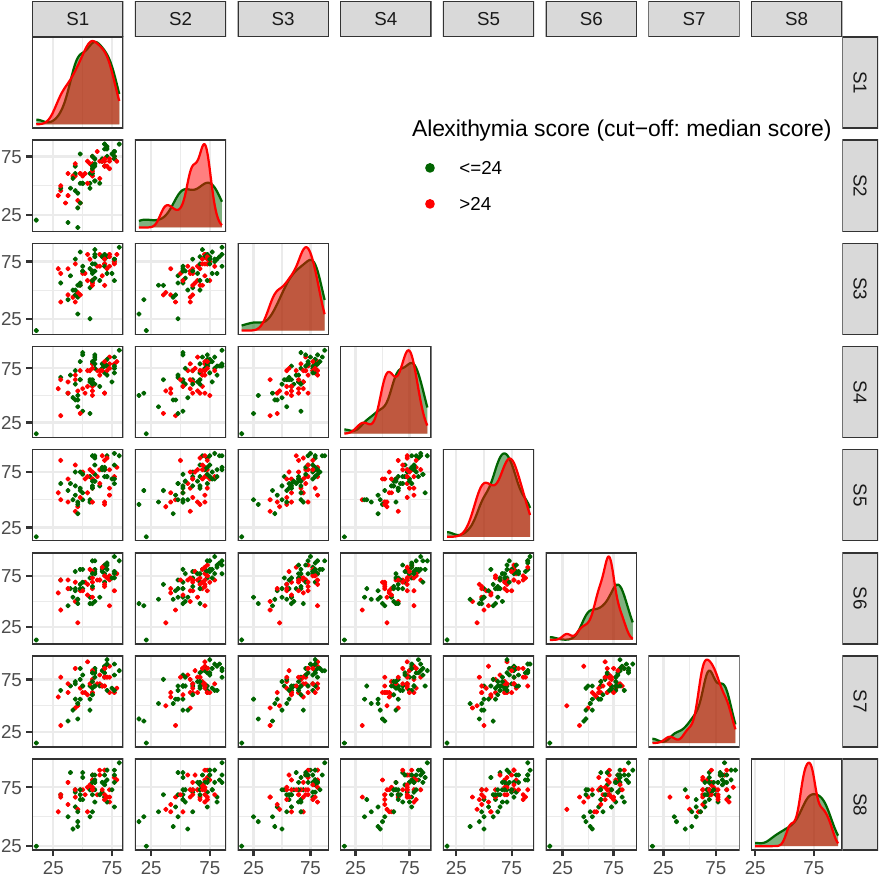

*Scatterplot matrix of accuracy score by alexithymia score for all participants (n=88). The density plots of accuracy scores at corresponding sessions are depicted on the diagonal for each level of the distinguishing factor.*

### Figure S14. Scatterplot matrix of accuracy score across all pairs of sessions by learning disability diagnosis

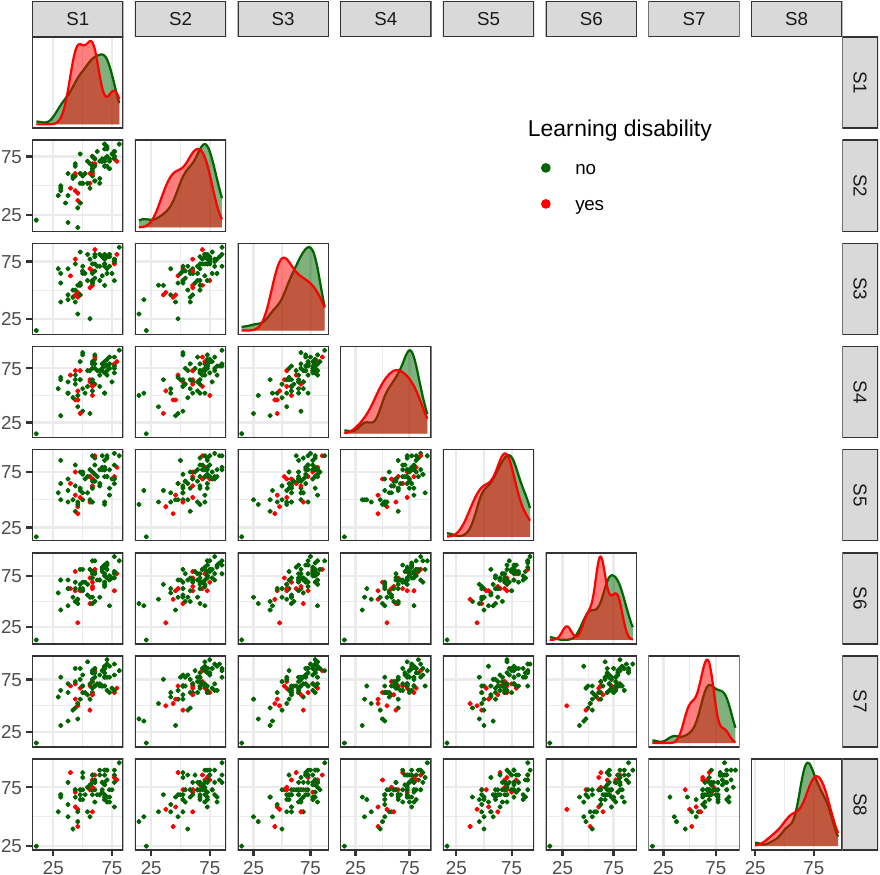

*Scatterplot matrix of accuracy score by learning disability diagnosis for all participants (n=88). The density plots of accuracy scores at corresponding sessions are depicted on the diagonal for each level of the distinguishing factor.*

### Figure S15. Scatterplot matrix of accuracy score across all pairs of sessions by ADHD diagnosis

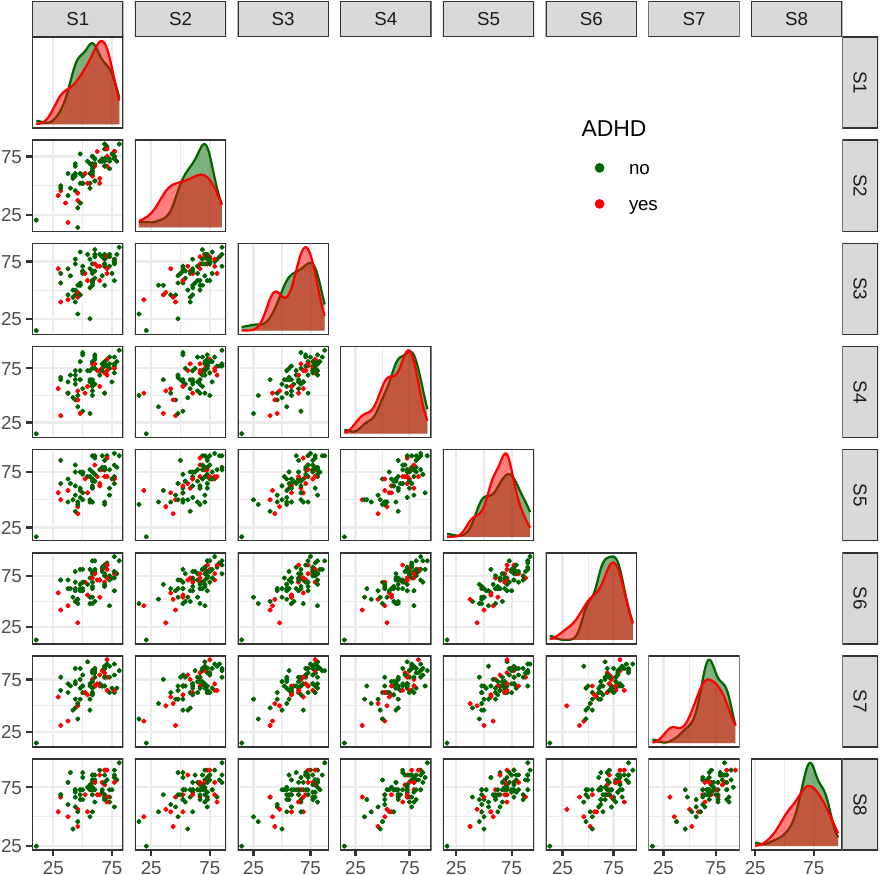

*Scatterplot matrix of accuracy score by ADHD diagnosis for all participants (n=88). The density plots of accuracy scores at corresponding sessions are depicted on the diagonal for each level of the distinguishing factor.*

### Figure S16. Scatterplot matrix of accuracy score across all pairs of sessions by dyslexia diagnosis

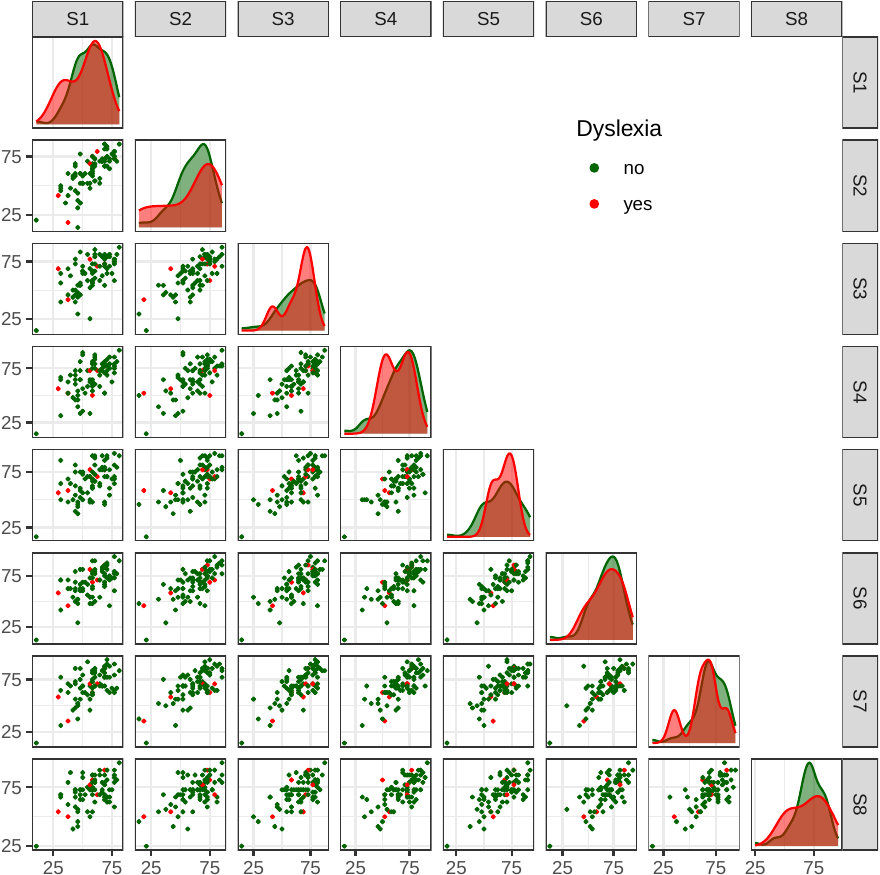

*Scatterplot matrix of accuracy score by dyslexia diagnosis for all participants (n=88). The density plots of accuracy scores at corresponding sessions are depicted on the diagonal for each level of the distinguishing factor.*

### Figure S17. Scatterplot matrix of accuracy score across all pairs of sessions by dyspraxia diagnosis

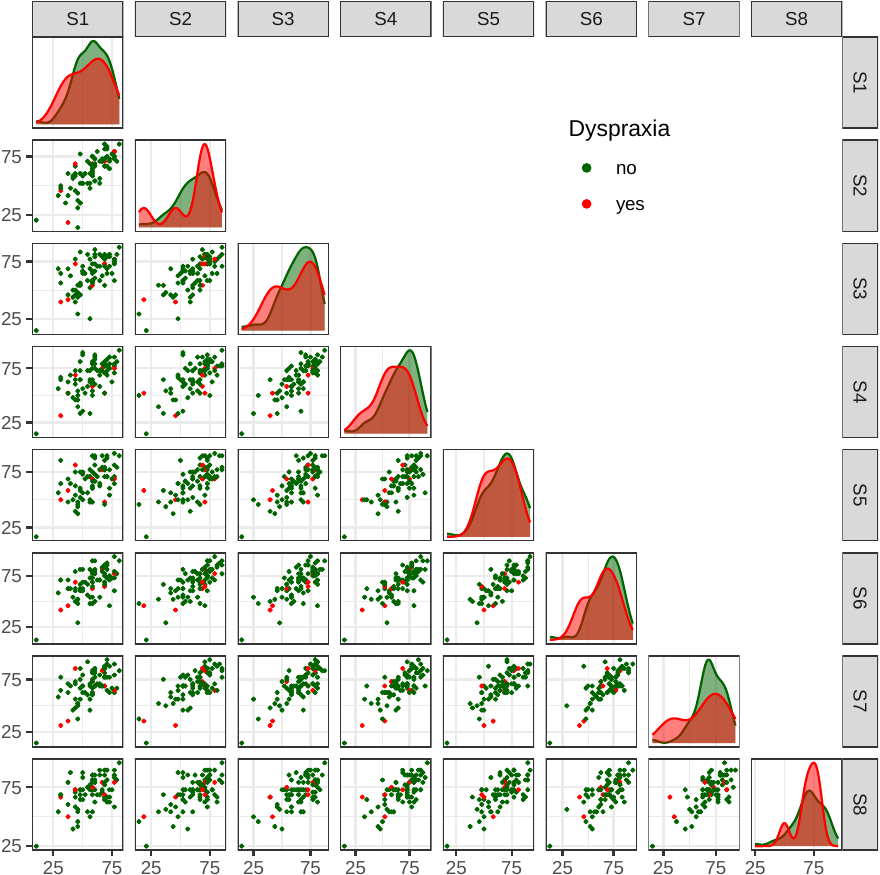

*Scatterplot matrix of accuracy score by dyspraxia diagnosis for all participants (n=88). The density plots of accuracy scores at corresponding sessions are depicted on the diagonal for each level of the distinguishing factor.*

### Figure S18. Box-plots of mean number of attempts across sessions for all participants (n=88)

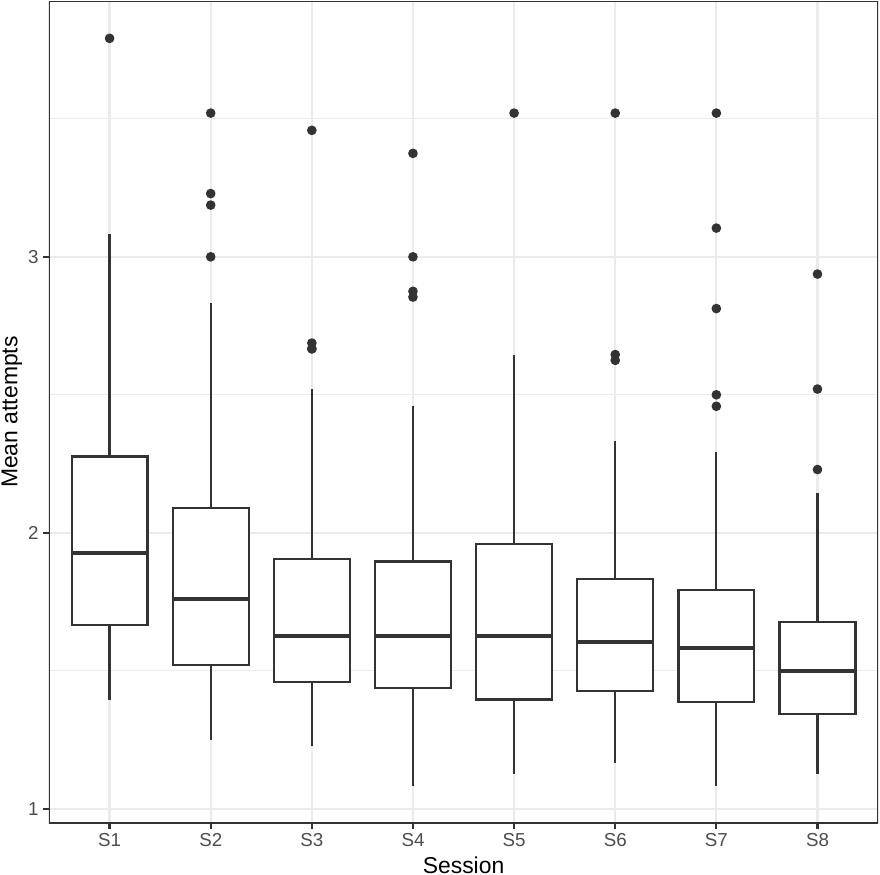

### Figure S19. Box-plots of false alarms per emotion across sessions for all participants (n=88)

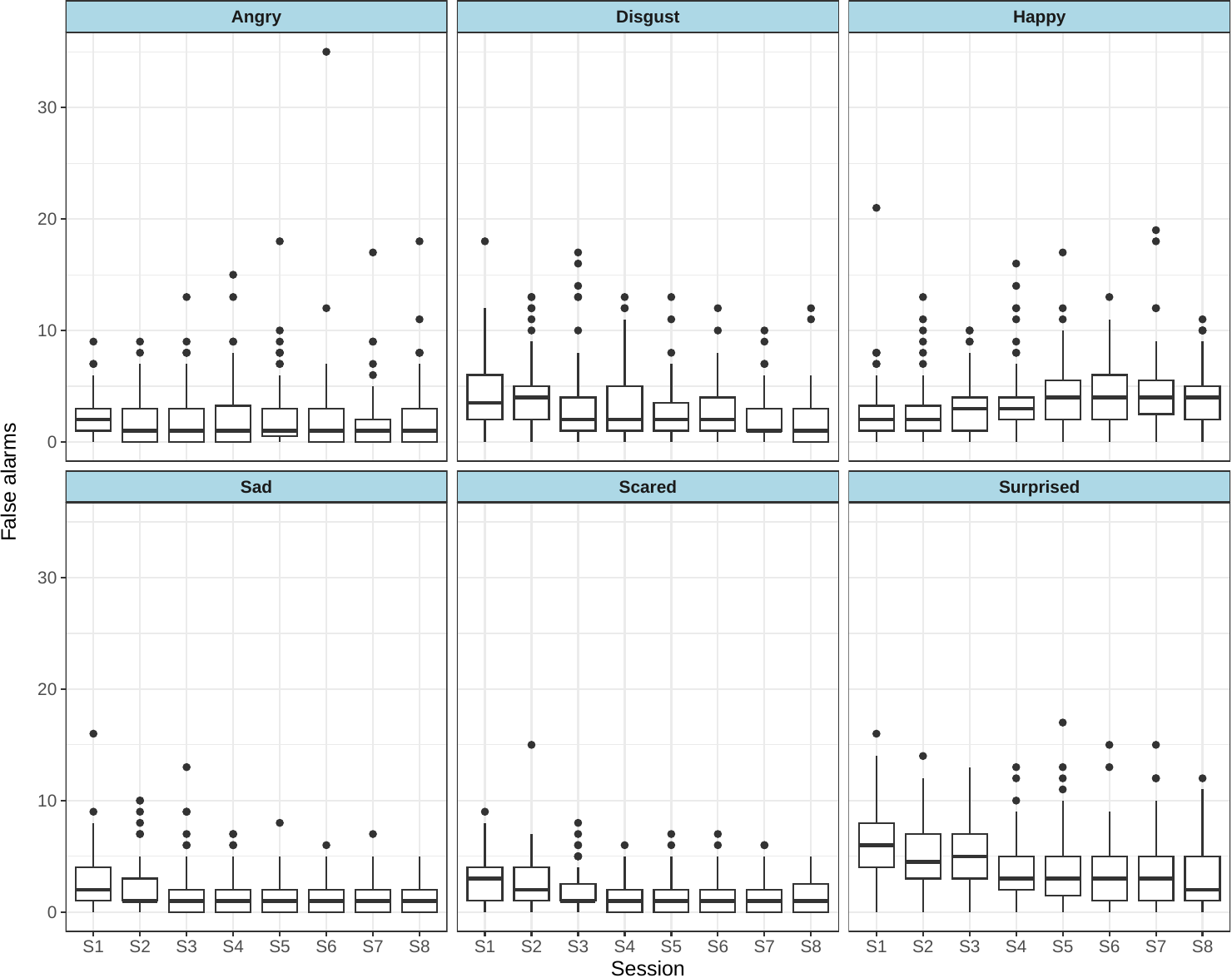

### Figure S20. Box-plots of A-prime sensitivity scores per emotion across sessions for all participants (n=88)

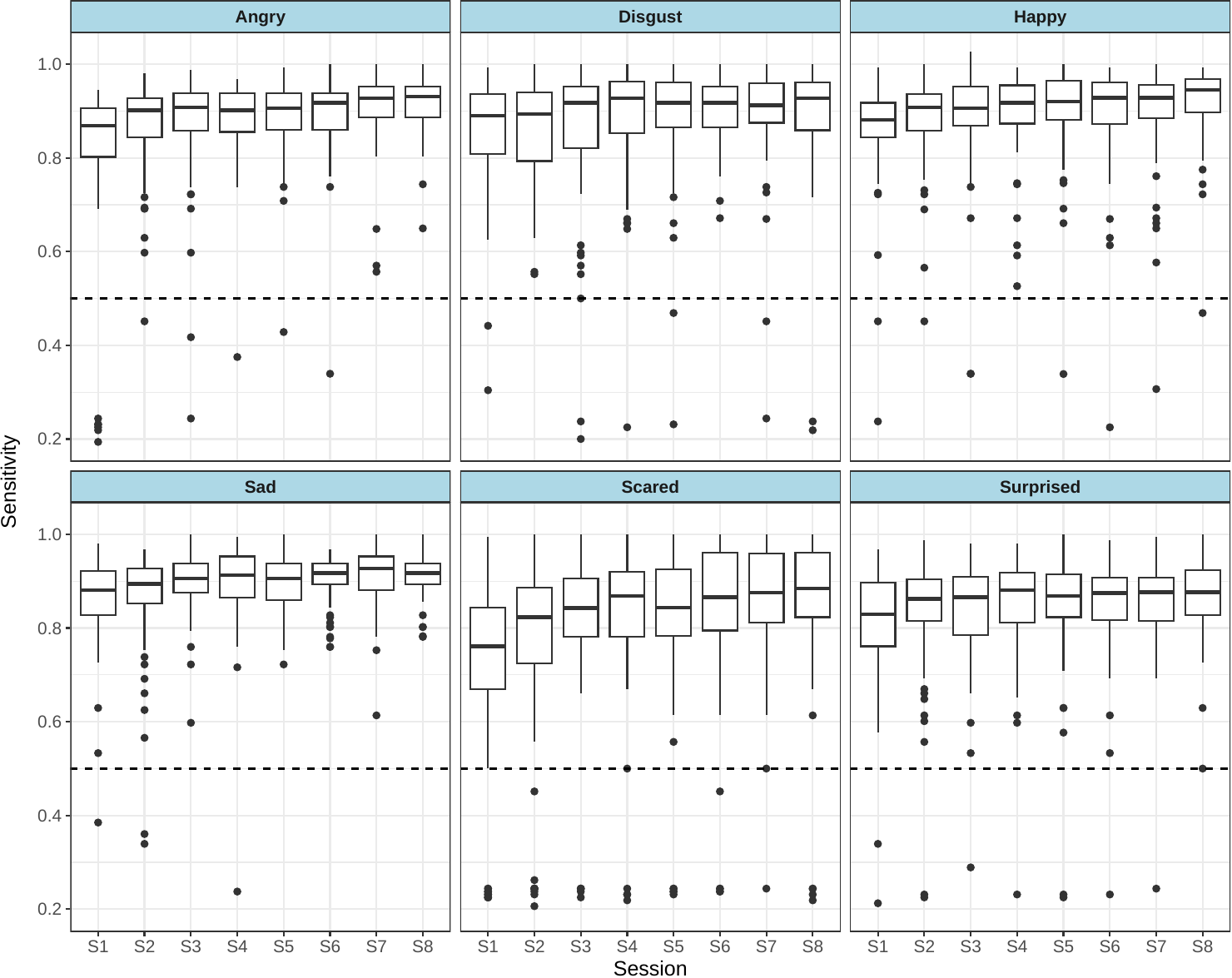

*The level of random guess (A-prime score of 0.5) has been represented by the dashed horizontal lines.*
